# Severe Neuro-COVID is associated with peripheral immune signatures, autoimmunity and signs of neurodegeneration: a prospective cross-sectional study

**DOI:** 10.1101/2022.02.18.22271039

**Authors:** Manina M. Etter, Tomás A. Martins, Laila Kulsvehagen, Elisabeth Pössnecker, Wandrille Duchemin, Sabrina Hogan, Gretel Sanabria-Diaz, Jannis Müller, Alessio Chiappini, Jonathan Rychen, Noëmi Eberhard, Lester Melie-Garcia, Emanuela Keller, Ilijas Jelcic, Hans Pargger, Martin Siegemund, Jens Kuhle, Johanna Oechtering, Caroline Eich, Alexandar Tzankov, Matthias S. Matter, Özgür Yaldizli, Johanna M. Lieb, Marios-Nikos Psychogios, Caroline M. Berkemeier, Karoline Leuzinger, Hans H. Hirsch, Cristina Granziera, Anne-Katrin Pröbstel, Gregor Hutter

## Abstract

**Importance:** Growing evidence suggests that coronavirus disease 2019 (COVID-19) is associated with neurological sequelae. However, the underlying pathophysiological mechanisms resulting in central nervous system (CNS) derogation remain unclear.

**Objective:** To identify severity-dependent immune mechanisms in the cerebrospinal fluid (CSF) and plasma of COVID-19 patients and their association with brain imaging alterations.

**Design:** Prospective cross-sectional cohort study.

**Setting:** This study was performed from August 2020 to April 2021. Participants were enrolled in the outpatient clinics, hospital wards and intensive care units (ICU) of two clinical sites in Basel and Zurich, Switzerland.

**Participants:** Age >18 years and a positive SARS-CoV-2 test result were inclusion criteria. Potentially matching individuals were identified (n=310), of which 269 declined to participate and 1 did not match inclusion criteria. Paired CSF and plasma samples, as well as brain images, were acquired. The COVID-19 cohort (n=40; mean [SD] age, 54 [20] years; 17 women (42%)) was prospectively assorted by neurological symptom severity (classes I, II and III). Age/sex-matched inflammatory (n=25) and healthy (n=25) CSF and plasma control samples were obtained. For volumetric brain analysis, a healthy age/sex-matched control cohort (n=36) was established.

**Exposures:** Lumbar puncture, blood sampling and cranial MRI and/or CT.

**Main outcomes and measures:** Proteomics, standard parameters and antibody profiling of paired CSF and plasma samples in COVID-19 patients and controls. Brain imaging and gray matter volumetric analysis in association with biomarker profiles. Follow-up after 10-months.

**Results:** COVID-19 patients displayed a plasma cytokine storm but a non-inflammatory CSF profile. Class III patients displayed signs of blood-brain barrier (BBB) impairment and a polyclonal B cell response targeting self- and non-self antigens. Decreased regional brain volumes were present in COVID-19 patients and associated with specific CSF and plasma parameters.

**Conclusion and relevance:** Neuro-COVID class III patients had a strong, peripheral immune response resulting in (1) BBB impairment (2) ingress of (auto-)antibodies, (3) microglia activation and neuronal damage signatures. Our data point towards several potentially actionable targets that may be addressed to prevent COVID-19-related neurological sequelae.

**Trial registration:** The trial (NCT04472013) was registered on clinicaltrials.gov.

**Key points:** *Question:* Does a severity-dependent pattern of immune mechanisms exist in the cerebrospinal fluid (CSF) and plasma of COVID-19 patients and are these associated with clinical and brain imaging findings?

*Findings:* Neuro-COVID patients display a robust class III-specific peripheral immune response resulting in (1) blood-brain barrier (BBB) impairment, (2) ingress of (auto-)antibodies, (3) microglia activation and neuronal damage signatures. Integration of MRIs, brain volumetry and proteomics identified biomarkers associated with regional brain volume loss in severe Neuro-COVID.

*Meaning:* We provide a multidimensional framework of mechanisms associated with severe Neuro-COVID and present possible targets to prevent COVID-19-related neurological sequelae.

## Introduction

The prevalence of neurological symptoms after severe acute respiratory syndrome coronavirus 2 (SARS-CoV-2) infection, termed “Neuro-COVID”, differs significantly between studies and can rarely be explained by direct virus effects (1)(2).

Neuropathological evidence of hyperactive microglia supports detrimental immune responses in coronavirus disease 2019 (COVID-19) (3), and *post-mortem* studies postulate activated microglia as dominant immune cell population in COVID-19 brains. Additionally, formation of microglia and T cell nodules were detected across brain compartments as a site of greatest T cell and microglia activation (1).

Accordingly, cerebrospinal fluid (CSF) single-cell transcriptomics identified immune alterations in Neuro-COVID patients (2).

Schwabenland (1) confirmed the presence of amyloid precursor protein deposits in COVID-19 brains, suggesting axonal damage as a result of immune activation. However, SARS-CoV-2 RNA has rarely been detected in the CSF of COVID-19 patients, even in those displaying neurological symptoms. Moreover, new-onset humoral autoimmunity, including antineuronal antibodies, in COVID-19 individuals has been observed, even in the absence of increased conventional inflammatory CSF parameters and lacking evidence of inflammation upon neuroimaging (4)(5). Yet, it still remains controversial whether these alterations represent specific central nervous system (CNS) infection or are bystander effects of systemic COVID-19.

To understand the immune mechanisms responsible for different manifestations and severity classes of Neuro-COVID, we performed a prospective, in-depth characterization of immune mediators in the CSF and plasma of clinically well-characterized severity-stratified Neuro-COVID patients. Furthermore, we correlated these findings with imaging data, including magnetic resonance imaging (MRI), gray matter volume (GMV) and choroid plexus volume (CPV) analysis.

## Methods

### Study design, patient population and study interventions

We conducted a prospective, two-center, cross-sectional study (clinicaltrials.gov NCT04472013, IRB approval EKNZ 2020-01503) including COVID-19 patients from August 2020 to April 2021 at the Swiss University Hospitals of Basel and Zurich. Inclusion criteria were age >18 years and a real-time quantitative PCR (qRT-PCR)-positive SARS-CoV-2 infection. We screened 310 patients, of which 269 declined to participate and 1 did not meet inclusion criteria **(Figure 1A)**. The 40 enrolled patients underwent detailed neurological examination and were subdivided into three Neuro-COVID severity classes at the time of their positive SARS-CoV-2 qRT-PCR test, referred to as Neuro-COVID class I (n=18), II (n=7) or III (n=15). Class I was defined by mild (e.g. anosmia, ageusia, headache, dizziness), class II by moderate (e.g. mono/para/quadriparesis, fatigue) and class III by severe symptoms (e.g., strokes, seizures, cognitive impairment) (6).

**Figure 1:**
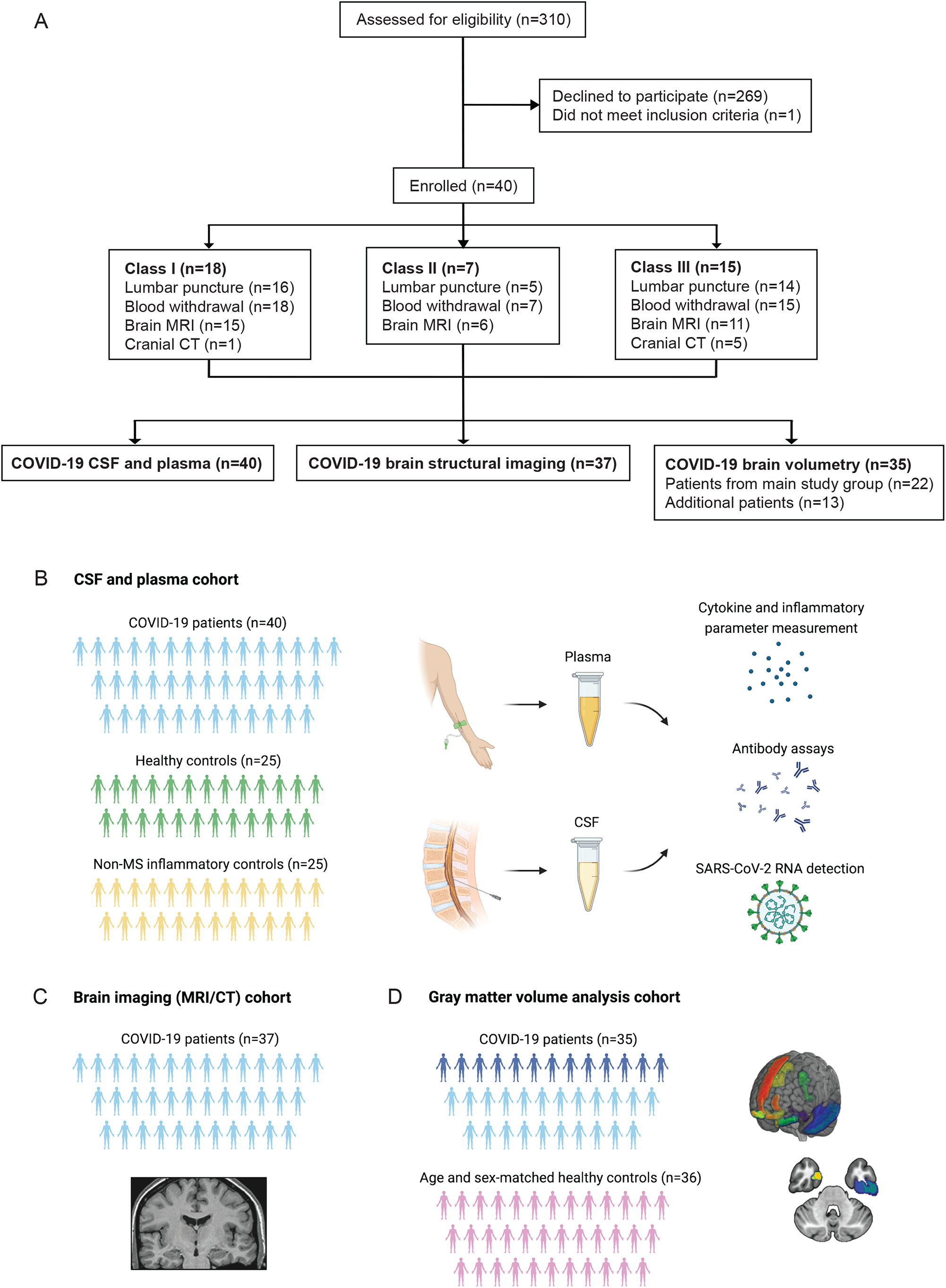
CONSORT diagram and schemes illustrating the project design. **A)** Consort flow diagram. Patients who tested positive for SARS-CoV-2 were assessed for eligibility (n=310), of which 269 declined to participate and 1 failed to meet inclusion criteria. Enrolled patients (n=40) were allocated to different severity classes of Neuro-COVID according to Fotuhi et. al. (6) with 18 in class I, 7 in class II and 15 in class III. *Schemes illustrating the study design:* **B)** Paired CSF and plasma samples were collected from 40 COVID-19 patients. Paired CSF and plasma samples from healthy and non-MS inflammatory neurological disease controls were retrospectively obtained. **C)** In 37 of the COVID-19 patients, a contrast-enhanced MRI or CT scan was conducted and evaluated by a board-certified neuroradiologist. **D)** Brain volumetric analysis was performed in 35 COVID-19 patients. This cohort included 22 patients of the main study cohort from whom Magnetization prepared - rapid gradient echo (MPRAGE) pulse sequences and paired CSF and plasma samples were obtained (*light blue*) and an additional 13 patients who underwent brain MRI during COVID-19 infection (*dark blue*). A cohort of 36 healthy age and sex-matched individuals served as the control group. **B-D)** Created with BioRender.com.

Study interventions included lumbar puncture and blood withdrawal for CSF/plasma soluble protein analysis **(Figure 1B, Supplementary methods)**, cranial MRI or CT and GMV and CPV analyses **(Figure 1C, 1D, Supplementary methods)**. Ten months after diagnosis, the COVID-19 Yorkshire rehabilitation scale (C19-YRS) recorded patients’ outcome **(Table 1)**.

**Table 1:** Characteristics of COVID-19 patients. Demographics, outcomes, clinical and paraclinical characteristics of different Neuro-COVID class patients. Patients could have more than 1 pre-existing illness in past medical history.

|  | **Class I (n=18)** | **Class II (n=7)** | **Class III (n=15)** |
| --- | --- | --- | --- |
| **Age, years, mean (SD)** | 48 (21) | 49 (19) | 62 (17) |
| Range, years | 22-80 | 23-73 | 22-98 |
| **Sex** |  |  |  |
| Female, n (%) | 8 (44.4%) | 3 (42.9%) | 6 (40.0%) |
| Male, n (%) | 10 (55.6%) | 4 (57.1%) | 9 (60.0%) |
| **Delay, d, mean (SD)** |  |  |  |
| Diagnosis to LP | 3 (3) | 3 (2) | 4 (4) |
| Range, d | (0-8) | (1-5) | (0-11) |
| Diagnosis to blood withdrawal | 3 (3) | 3 (2) | 4 (4) |
| Range, d | (0-8) | (0-7) | (0-15) |
| Diagnosis to MRI/CT | 4 (3) | 4 (4) | 6 (7) |
| Range, d | (1-11) | (0-11) | (0-12) |
| **Past medical history** |  |  |  |
| Arterial hypertension, n (%) | 4 (22.2%) | 1 (14.3%) | 7 (46.7%) |
| Type 2 diabetes, n (%) | 2 (11.1%) | 1 (14.3%) | 8 (53.3%) |
| Dyslipidemia, n (%) | 4 (22.2%) | 0 (0%) | 2 (13.3%) |
| Chronic kidney disease, n (%) | 2 (11.1%) | 0 (0%) | 4 (26.7%) |
| Coronary artery disease, n (%) | 4 (22.2%) | 1 (14.3%) | 1 (6.7%) |
| Cancer of any type, n (%) | 0 (0%) | 1 (14.3%) | 2 (13.3%) |
| COPD, n (%) | 0 (0%) | 0 (0%) | 0 (0%) |
| **Pre-existing neurological disorder** |  |  |  |
| Multiple sclerosis, n (%) | 1 (5.6%) | 0 (0%) | 0 (0%) |
| Underwent LP, n | 0 | - | - |
| Underwent blood withdrawal, n | 1 | - | - |
| Myasthenia gravis, n (%) | 0 (0%) | 1 (14.3%) | 0(0%) |
| Underwent LP, n | - | 0 | - |
| Underwent blood withdrawal | - | 1 | - |
| **Clinical evolution** |  |  |  |
| ICU, n (%) | 0 (0%) | 0 (0%) | 11 (73.3%) |
| Hospital ward, n (%) | 13 (72.2%) | 7 (100%) | 4 (26.7%) |
| Outpatient clinics, n (%) | 5 (27.8%) | 0 (0%) | 0 (0%) |
| **10-months follow-up** |  |  |  |
| Follow-up performed, n (%) | 18 (100%) | 7 (100%) | 8 (53.3%) |
| Lost to follow-up, n (%) | 0 (0%) | 0 (0%) | 7 (46.7%) |
| Full recovery without any new deficits, n (%)* | 6 (35.3%) | 1 (14.3%) | 1 (33.3%) |
| Concentration problems, n (%)* | 3 (17.6%) | 5 (71.4%) | 0 (0%) |
| Memory problems, n (%)* | 3 (17.6%) | 4 (57.1%) | 0 (0%) |
| Chronic fatigue, n (%)* | 0 (0%) | 5 (71.4%) | 2 (66.7%) |
| Speech/communication difficulties, n (%)* | 0 (0%) | 1 (14.3%) | 0 (0%) |
| Change/loss of smell, n (%)* | 0 (0%) | 2 (28.6%) | 0 (0%) |
| Change/loss of taste, n (%)* | 1 (5.9%) | 1 (14.3%) | 0 (0%) |
| Muscle pain, n (%)* | 2 (11.8%) | 1 (14.3%) | 0 (0%) |
| Difficulties in daily activities, n (%)* | 3 (17.6%) | 2 (28.6%) | 1 (33.3%) |
| Deceased, n (%) | 1 (5.6%) | 0 (0%) | 5 (62.5%) |
| In hospital during the ongoing study, n (%)* | 0 (0%) | - | 3 (37.5%) |
| **CSF characteristics** |  |  |  |
| **Protein levels** |  |  |  |
| Missing, n | 4 | 2 | 1 |
| Protein levels, mg/L, mean (SD) | 334 (124) | 282 (114) | 1424 (3056) |
| Range, mg/L | 103-673 | 174-429 | 286-12000 |
| Elevated protein (>500mg/L), n (%) | 1 (7.1%) | 0 (0%) | 10 (71.4%) |
| **Leukocyte count** |  |  |  |
| Missing, n | 5 | 2 | 2 |
| Leukocyte count, x10<sup>6</sup>/L, mean (SD) | 3 (3) | 3 (2) | 12 (27) |
| Range, x10<sup>6</sup>/L | 1-12 | 1-7 | 0-99 |
| Elevated leukocyte count (≥5x10<sup>6</sup>/L), n (%) | 1 (7.7%) | 1 (20%) | 6 (46.2%) |
| **Albumin ratio (CSF/plasma)** |  |  |  |
| Missing, n | 6 | 4 | 6 |
| Ratio, mean (SD) | 5.2 (2.1) | 5.2 (3.1) | 13.1 (7.8) |
| Elevated ratio (≥8.2x10<sup>-3</sup>), n (%) | 1 (8.3%) | 1 (33.3%) | 6 (66.7%) |
| **Glucose levels** |  |  |  |
| Missing, n | 4 | 2 | 2 |
| Glucose levels, mmol/L, mean (SD) | 4.3 (1.2) | 4.8 (1.5) | 5.3 (1.8) |
| Range, mmol/L | 3.2-7.6 | 3.5-7.2 | 2.9-7.9 |
| Elevated glucose (>6.1mmol/L), n (%) | 1 (7.1%) | 1 (20%) | 4 (30.8%) |
| **Lactate levels** |  |  |  |
| Missing, n | 4 | 2 | 3 |
| Lactate levels, mmol/L, mean (SD) | 1.7 (0.3) | 2.0 (0.7) | 2.8 (1.9) |
| Range, mmol/L | 1.4-2.2 | 1.3-2.9 | 1.6-8.4 |
| Elevated lactate (>2mmol/L), n (%) | 3 (21.4%) | 2 (40%) | 9 (75%) |
| SD: standard deviation<br>LP: lumbar puncture<br>MRI: magnetic resonance imaging<br>CT: computed tomography<br>COPD: chronic obstructive pulmonary disease<br>CSF: cerebrospinal fluid<br>*percentage value refers to the number of conducted follow-ups |  |  |  |

Retrospectively, biobanked age- and sex-matched CSF/plasma samples from patients with non-MS inflammatory neurologic disorders (n=25, IC) and healthy individuals (n=25, HC) served as controls **(Table S1)**.

### Statistical analysis of CSF and plasma proteomics

Following imputation, normalized protein expression values (NPX) were adjusted on the median age and sex. Kendall’s Tau method correlated CSF/plasma expression levels. Group comparison was carried out using the Mann-Whitney-U test with a Benjamin-Hochberg (BH) correction for multiple testing.

### Statistical analysis of volumetric brain imaging

Using the Shapiro-Wilk test normal distributions of variables were confirmed. Equal variance between groups was assessed by Levene’s test. Clinical and demographic variables were compared with independent t-test, Mann-Whitney-U test, or Chi-square test. Regional volumes were compared using a linear regression model. Additional covariates were age, sex, age*sex interaction, MRI magnetic field strength and total intracranial volume (TIV). Choroid plexus volume adjusted for TIV was compared by Mann-Whitney-U test. Associations between GMV and clinical measures were assessed using partial correlations. P-values were adjusted for multiple comparisons using false discovery rate (FDR). The analysis was performed using JASP (https://jasp-stats.org/) and MATLAB (‘partialcorri.m’ function) (https://www.mathworks.com/). Further details are described in **Supplementary methods**.

## Results

### Characteristics of the study cohort and follow-up

Forty patients with qRT-PCR-confirmed SARS-CoV-2 infection (mean [SD] age, 54 [20] years; 17 women [42%]) were enrolled in this cross-sectional analysis using a prospective study design **(Figure 1A)**. Patient characteristics and follow-up details per severity class are summarized in **Table 1**. COVID-19 patients were classified into absent/mild (n=18), moderate (n=7) and severe (n=15) Neuro-COVID classes I, II and III **(Figure 1A, Table 1, Table S2)**. Study interventions are summarized in **Figure 1B-D**. The results of the 10-months follow-up are described in **Table 1**. Our long-term follow-up suggests that class II and III patients continued to be more often affected by neurological problems compared to class I. In class I, one third recovered without any long-term deficits. Notably, the mortality rate was high in class III patients: Five deceased, 3 of them during their hospital stay and 2 during the follow-up period.

### Class III patients have an impaired blood-brain barrier (BBB) and a polyclonal B cell response

In class III patients, CSF protein and lactate levels were significantly increased compared to class I/II **(Figure 2A)**. In contrast, CSF leukocytes were not elevated in COVID-19 patients, which signified ICs. Importantly, CSF glucose was increased even in class I and II patients versus (vs) HCs and was a significant discriminator between class III and ICs **(Figure 2A)**. Routine CSF parameters are described in **Table 1** and **Table S3**. In tendency, the CSF/plasma albumin ratio **(Figure 2A)** as well as the total CSF IgG **(Figure S1B)** were higher in class III patients. In line with recent research (2), SARS-CoV-2 RNA was not detectable in the CSF. However, we were able to detect SARS-CoV-2 Spike (S) protein antibodies in 12 plasma and 3 CSF samples **(Figure 2C)**, yet the antibody index (AI) pointed to a peripheral synthesis of these intrathecal antibodies **(Table S3)**. We could not detect reactivities against known CNS myelin antigens **(Figure S1**, CSF data not shown), but found elevated anti-dsDNA-IgG/IgA and anti-gut microbiota IgA responses in the CSF of class III compared to class I patients and ICs **(Figure 2C, Figure S1)**. This was paralleled by an elevated anti-BSA reactivity. The AI pointed towards a peripheral production of these polyclonal antibodies.

**Figure 2:**
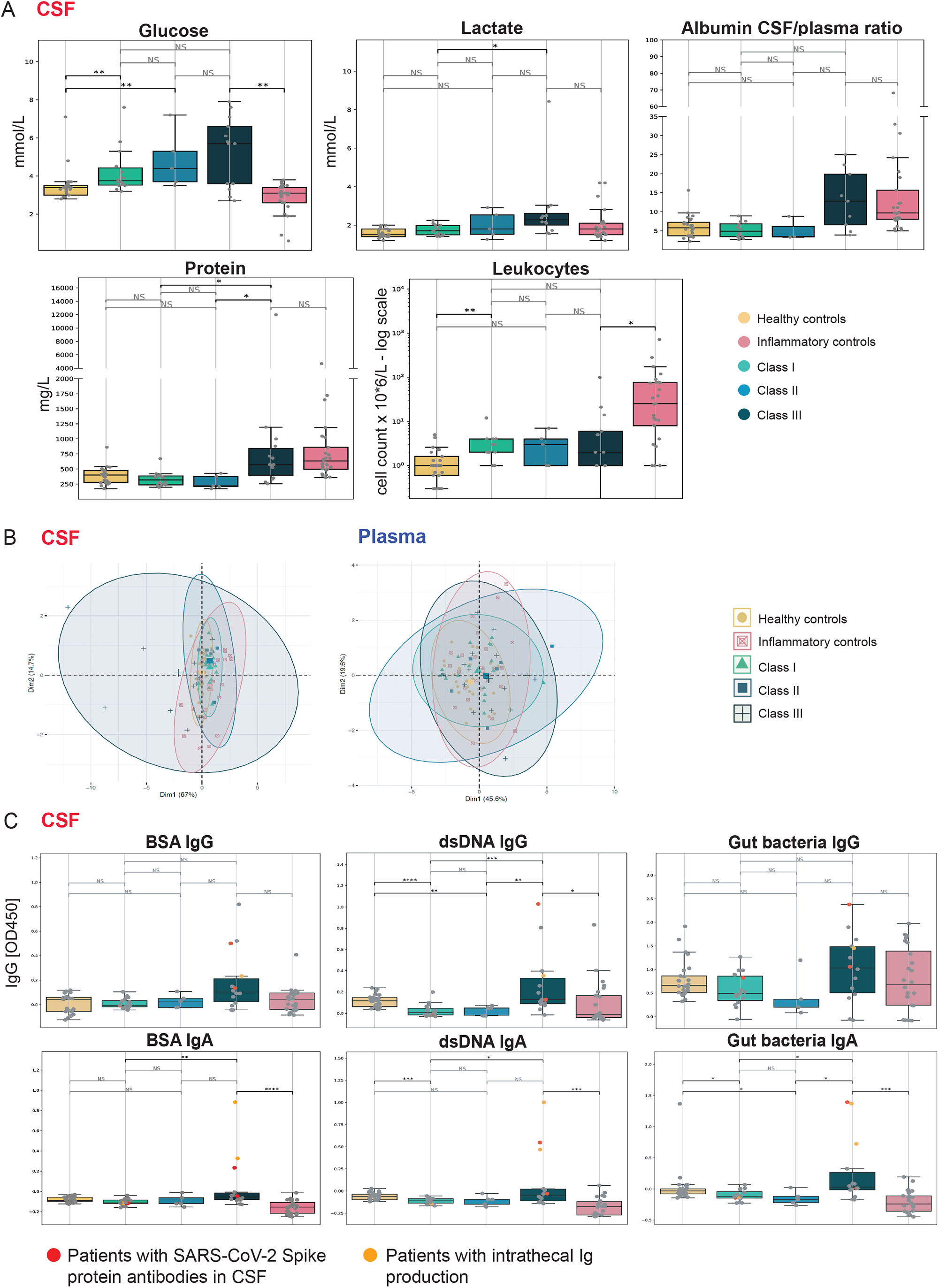
Routine inflammatory CSF parameters and B cell response in Neuro-COVID patients. **A)** Box plot representation of routine CSF parameters including glucose (mmol/L), lactate (mmol/L), albumin CSF/plasma ratio, total protein (mg/L) and leukocytes (cell count × 10^6^/L - log scale). Statistics: The data for each parameter, except the leukocyte count, was marginalized on sex and age. **B)** Principal component analysis (PCA) plot of CSF *(left plot)* and plasma *(right plot)* BSA IgG and IgA, dsDNA IgG and IgA, RePOOPulate IgG and IgA. The CSF antibody signature of class III predominantly differs from inflammatory and HCs. Dim1: first principal component; Dim2: second principal component; Percentages account for total variance of the data; Data points colored by category; Bigger points represent centroids. **C)** Box plot representation of CSF levels (OD450; optical density at 450 nm) of anti-BSA, anti-dsDNA-and anti-gut microbiota (RePOOPulate)-IgG/IgA per patient and control group. Patients with SARS-CoV-2 S Protein antibodies in the CSF indicated in red, those with intrathecal IgG or IgA production in orange, respectively. Mann-Whitney-U test (NS: not significant, adjusted p: *<0.05, **<0.01, ***<0.001).

### Targeted proteomic analysis of CSF and plasma reveals a vigorous peripheral immune response in Neuro-COVID and a class III-specific signature

We identified predominant plasma secretion of a large number of soluble proteins in Neuro-COVID class III patients compared to controls **(Figure 3A, Figure S2A)**, suggesting a Neuro-COVID class-dependent plasma signature. Class I and II patients had an increased plasma secretome compared to controls, in line with the previously described peripheral cytokine storm in COVID-19 (7).

**Figure 3:**
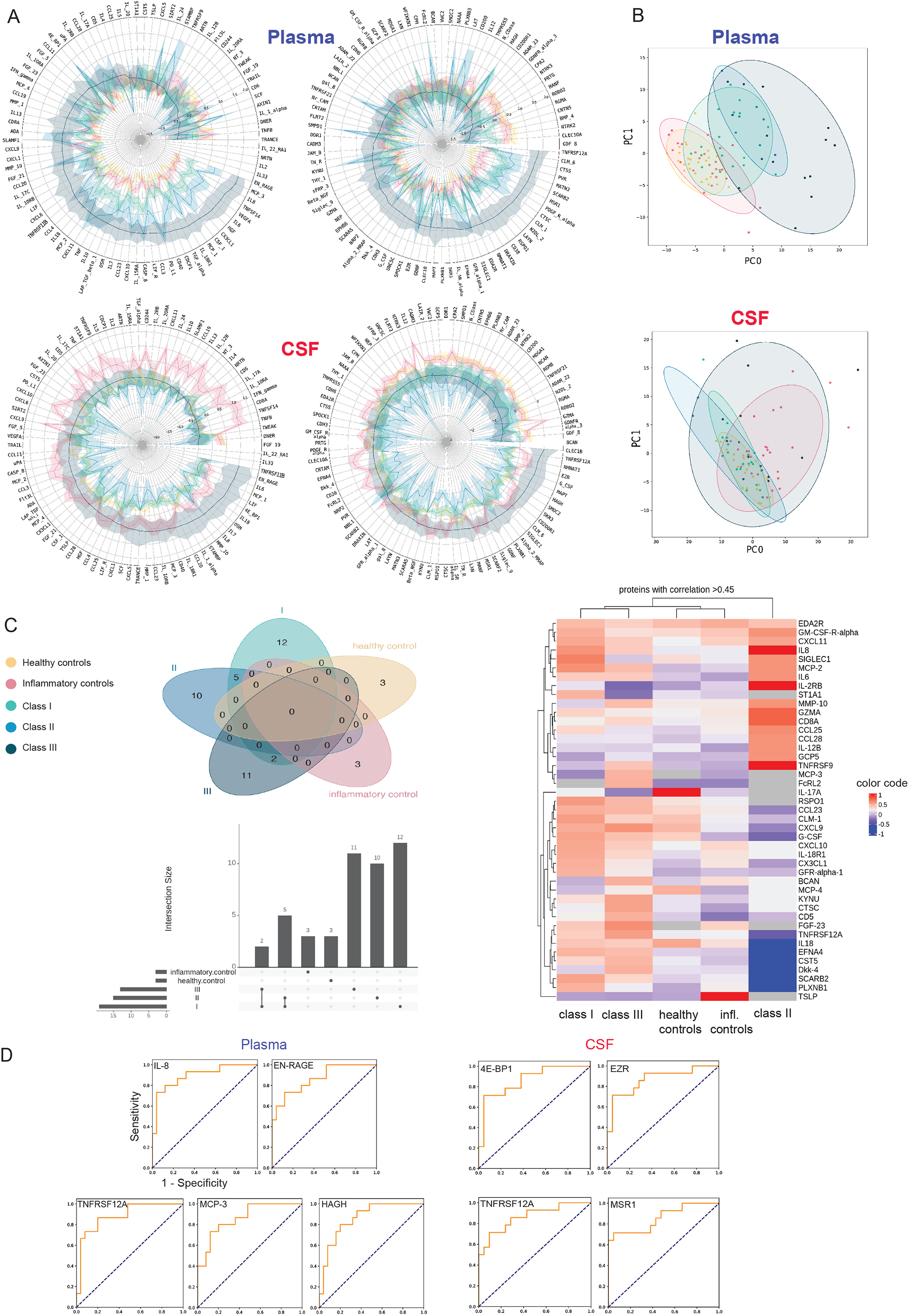
Neuro-COVID patients display a vigorous peripheral immune response including analytes with high predictive value for class III development and strong CSF-plasma correlation. **A)** Rose plots representing Z-scores of marginalized NPX of 192 soluble proteins in CSF and plasma. For better visualization, analytes have been grouped into ‘inflammatory’ *(left panels)* and ‘neurological’ *(right panels)* proteins. **B)** Principal component analysis (PCA) of 192 examined soluble proteins in CSF and plasma. Each patient is presented by one dot, and colored according to healthy/IC or COVID-class I-III. The ellipses represent the 95% confidence interval within subgroups. PC0 and PC1 components are listed in **Table S10. C)** CSF-plasma correlation analysis with Venn diagram, UpSet plot and a heatmap of the strongest correlated genes (correlation coefficient >0.45). Venn diagram and UpSet plot demonstrate 10-12 class-defining proteins with a strong CSF-plasma correlation and only a few overlapping proteins. CSF-plasma correlation values are color-coded, indicating strong correlation values in red color and low correlation values in blue color. Heatmap depicts a myeloid/eosinophil proinflammatory signature in class I patients, changing to a T cell-mediated, proinflammatory feature in class II patients. Myeloid signature correlations are preserved in class II and partly overlap with class I patients. In class III the strongest CSF-plasma correlation pattern is characterized by biomarkers implicating tissue damage and neuronal damage. **D)** ROC-AUC analysis of class I and II vs class III. Five predictive plasma markers, including IL-8, EN-RAGE, TNFRSF12A, MCP-3, and 4 CSF markers, including 4E-BP1, EZR, TNFRSF12A, MSR1, emerged for the prediction of class III development. The Y-axis represents the sensitivity, the X-axis represents the 1 - specificity (represented for IL-8, plasma). The names of relevant proteins in the study are compiled in **Table S6**.

The CSF soluble protein pattern was different: while class I and II patients had relatively similar profiles as HC patients, a Neuro-COVID class III-specific signature with differences to ICs emerged **(Figure 3A, Figure S2B)**. Notably, CSF total protein levels progressively increased from class I to III, indicating a correlation between CSF proteomics and neurological symptoms **(Figure 3A)**.

Principal component analysis (PCA) of soluble proteins distinctly separated controls from COVID-19 patients. Protein patterns largely overlapped between HCs and ICs, whereas COVID-19 classes clustered apart from the controls, with class III patients displaying the strongest separation **(Figure 3B, Table S4)**.

Most proteins were intrathecally (CSF/plasma ratio >0) secreted in ICs, whereas proteins in COVID-19 patients were peripherally synthesized (CSF/plasma ratio <0). Of note, TRANCE/RANKL was the only intrathecally synthesized protein in class III compared to ICs **(Figure S3, Table S5, Table S6)**.

### Neuro-COVID class III features are manifestations of microglia regulation, neurodegeneration and BBB disruption

As reported previously, plasma IL-6, IL-8, EN-RAGE, HGF, VEGFA, PD-L1 and TNFRSF12A levels were associated with COVID-19 severity (8) and distinct from ICs **(Figure S4A)**, which lacked peripheral inflammation. Plasma TNFRSF11B, EZR and CCL23 were increased in class III vs ICs **(Figure S4A)**. In contrast, plasma BMP-4, CLEC10A, CNTN5, GDF-8, NTRK2, ROBO2 and GDNFRα3 levels were lower in class III compared to ICs and class I patients **(Figure S4B)** (9–16). Further, compared to ICs, class III patients displayed higher plasma 4E-BP1 **(Figure S4C)**. HAGH was the only protein displaying higher plasma levels in class I vs class III **(Figure S4D)**.

Several CSF soluble protein levels, particularly mediators of microglia regulation and neurodegeneration, including IL-8, MSR1, 4E-BP1, CD200R1, TNFRSF12A and EZR were increased in class III compared to class I **(Figure S4E)** (9)(10). However, only TNFRSF11B levels were both discriminating class III from ICs and gradually increasing among Neuro-COVID classes **(Figure S4F)**.

CSF-plasma correlations identify a neuronal damage signature in class III, encompassing predictive markers for severe Neuro-COVID Assuming a cut-off of >0.45 in the Kendall-Tau correlation matrix, class-specific CSF-plasma correlations were noted and ranked **(Figure 3C**, *Venn diagram*). ICs and HCs were characterized only by few strong correlations compared to the Neuro-COVID groups.

We observed a gradual change in correlations from class I to class III. Only a few overlapping soluble proteins with strong correlations were detected, whereas 10-12 individual class-defining proteins were identified **(Figure 3C**, *Venn diagram and UpSet plot*). In class I, the strongest correlations (value >0.55) were characterized by a myeloid signature: SIGLEC1, MCP2, IL-8 and CLM1 (11–14) **(Figure 3C**, *heatmap*). In class II, a T cell-mediated signature prevailed by CCL25, CD8A, GZMA, TNFRSF9 and IL2-RB, while some myeloid correlations overlapped between class I and II (15–17). In class III, the pattern shifted to a neuronal damage signature encompassing CTSC, KYNU, TNFRSF12A, and CXCL9 (9,10,18,19).

To forecast severe Neuro-COVID, we attempted to identify biomarkers displaying strong correlations. Nine analytes (4 CSF and 5 plasma proteins) displayed an AUC-ROC score of >0.85, suggesting a high predictive power for class III development **(Figure 3D, Table S7)**. Among these, TNFRSF12A had a strong correlation (class III: 0.56, class II: -0.4, class I: 0.2), validating it as a predictive biomarker for severe Neuro-COVID.

### Neuro-COVID class III patients feature striking findings on brain imaging while most class I and II patients lack evidence of neuroinflammation

Brain images of each class are depicted in **Figure 4A-C**. Detailed imaging findings are presented in **Table S8**. The most frequent MRI findings were bilateral, multifocal hyperintense signal abnormalities on fluid-attenuated inversion recovery (FLAIR)/T2-weighted (T2w) imaging (n=18, 56.3%; class I: n=5, 33.4%; class II: n=5, 83.4%; class III: n=11, 72.7%). These signal abnormalities were predominantly located in the periventricular region (13 patients, 40.6%) and the semioval center (16 patients, 50%). Further, diffusion-weighted imaging (DWI) changes were present in 4 patients (12.5%): 1 class I/II, and 2 class III patients. Black blood and/or time of flight (TOF) imaging was acquired in 4 patients, in 2 of which (both from class III) focal vessel wall enhancement was visible, indicative of cerebral vasculitis **(Figure 4C)**. No signal changes were detected in the olfactory bulb. In 3 CT-scanned class III patients, we found 1 infratentorial or supratentorial infarction, 1 thrombosis of the sigmoid sinus with intracerebral hemorrhage and 1 bifrontal subarachnoid hemorrhage **(Figure 4C)**.

**Figure 4:**
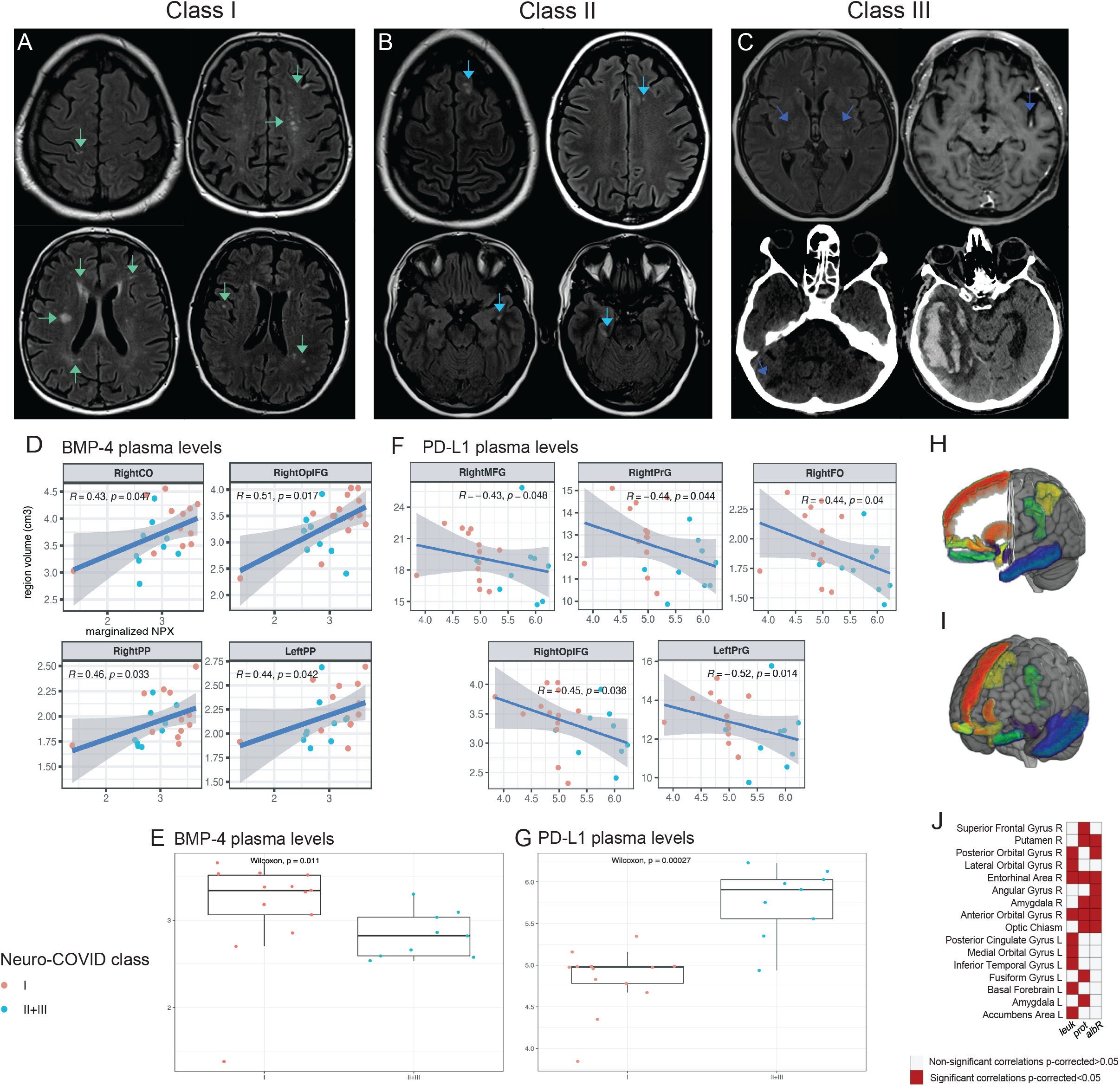
Routine brain imaging, regional GMV and association with different CSF and plasma analytes. **A-C)** Conventional brain MRI and CT scan analysis: exemplary imaging findings for each Neuro-COVID class. From left to right: class I **(A)**, class II **(B)**, class III **(C). A)** Class I: Axial FLAIR images of the same class I patient show multifocal hyperintensities in the right precentral gyrus (*top left*), semioval center and left frontal cortex *(top right)*, deep white matter and periventricular region *(bottom left)*, right temporal lobe and left parietal white matter *(bottom right)*. **B)** Class II: Axial FLAIR images of the same class II patient depict multifocal hyperintensities in the left frontal superior gyrus *(top left)*, white matter of left frontal lobe *(top right)*, left parahippocampal white matter *(bottom left)* and the right mesial temporal region *(bottom right)*. **C)** Class III: Axial FLAIR image shows bilateral thalamic hyperintensities *(top left)*. Axial T1-weighted (T1w)+ image depicts left middle cerebral artery (MCA) (M2-segment) enhancement in the insular cistern *(top right)*. Coronal cranial CT scan demonstrates right cerebellar infarction *(bottom left)*. Coronal cranial CT scan shows right temporo-occipital intracerebral hemorrhage *(bottom right)* (secondary to thrombosis of the right sigmoid sinus (not shown)). **D)** Correlation plots demonstrating the regional brain volumes significantly correlating with BMP-4 plasma levels in Neuro-COVID patients. The Y-axis represents the regional brain volume values. The X-axis represents the marginalized NPX of the respective protein. None of the adjusted p-values was significant after BH-procedure. **E)** Boxplot representation of marginalized NPX of class I compared to merged class II+II plasma BMP-4 (Wilcoxon, p=0.011). **F)** Correlation plots demonstrating the regional brain volumes significantly correlating with PD-L1 plasma levels in Neuro-COVID patients. The Y-axis represents the regional brain volume values. The X-axis represents the marginalized NPX of the respective protein. None of the adjusted p-values was significant after BH-procedure. **G)** Boxplot representation of marginalized NPX of class I compared to merged class II+II plasma PD-L1 (Wilcoxon, p=0.011). Panel **H)** and **I)** show the 3D view of the 16 brain regions with significant correlation values of GMV and clinical variables in the Neuro-COVID group after multiple comparison correction (FDR) These regions are represented in different colors on a T1w template (in radiological convention). Panel **J)** shows a matrix representing the association significance (significant p-corrected <0.05 in red squares). MRIcroGL software was used to generate this figure (https://www.nitrc.org/projects/mricrogl). Legend: RightCO: right central operculum, RightOpIFG: right opercular part of the inferior frontal gyrus, RightPP: right planum polare, LeftPP: left planum polare, RightMGF: right middle frontal gyrus, RightFO: right frontal operculum, RightPrG: right precentral gyrus, LeftPrG: left precentral gyrus, leuk: leukocytes, prot: protein, albR: Albumin CSF-plasma ratio.

### GMV in olfactory pathway structures decrease in Neuro-COVID patients and negatively correlate with inflammatory CSF parameters

Twenty brain regions displayed a smaller volume in the Neuro-COVID group compared to age/sex-matched HCs **(Table S9)**. Thereof, 81% corresponded to the olfactory/gustatory cortex’s telencephalic connections, including the amygdala, entorhinal cortex, basal ganglia cingulate gyrus and orbitofrontal areas. However, this finding was not significant after FDR correction. Further, we found negative correlations between regional GMVs and CSF leukocyte count, protein levels and CSF/plasma albumin ratio **(Table S10)** in the Neuro-COVID group within 16 specific brain regions **(Figure S5)**. CPVs did not differ significantly between COVID-19 patients and controls, but we found marginally higher CPVs in class III compared to class I/II. However, these differences were not significant (data not shown).

### PD-L1 and HGF plasma levels are associated with decreased regional GMV, while GDF-8 and BMP-4 are neuroprotective in Neuro-COVID patients

Additionally, we investigated the correlation of class-defining soluble CSF/plasma proteins and regional GMVs in regions with significantly lower volumes in COVID-19 patients **(Table S9)**. Plasma GDF-8 and BMP-4, implicated in neuroprotection and tissue reparatory responses (20,21) were significantly lower in class III and II compared to class I **(Figure 4D, 4E, Figure S6)**. Conversely, PD-L1 and HGF were associated with decreased GMVs in Neuro-COVID patients **(Figure 4F, Figure S6)**. PD-L1 and HGF plasma levels were higher in class III and II than in class I patients **(Figure 4G, Figure S6)**. Additional CSF and plasma proteins associated with particularly decreased and preserved GMVs underline the potential importance of COVID-19 dysregulated plasma and CSF proteins on brain structural changes **(Figure S7)**.

## Discussion

We identified Neuro-COVID-specific CSF and plasma alterations, providing insights into pathomechanisms underlying COVID-19-associated neurological sequelae. Compared to previous analyses, we studied the associations of peripheral inflammation, neuroinflammation and neurological symptoms multidimensionally within prospectively stratified Neuro-COVID classes. Remarkably, our long-term follow-up suggests that class II and III patients continued to be severely affected by neurological problems, and mortality was high in class III patients.

In line with other studies (22), we found elevated CSF glucose and lactate levels in class III patients. Indeed, patients suffering from diabetes were more prevalent in class III **(Table 1)**. The elevated lactate levels in class III potentially hint at cerebral hypoxia. However, class III patients had putative COVID-19-induced stroke or intracerebral hemorrhage, which may explain this finding.

Notably, we identified a class III-specific humoral CSF immune response encompassing enrichment of (total) IgG/IgA against self-(dsDNA) and non-self (BSA) antigens. This finding is corroborated by distinct plasma B cell clusters and antibody reactivity profiles previously reported (5). Certainly, antibody production predominantly took place in the plasma, pointing to an ingress of peripherally activated B cells/antibodies. In line with recent findings identifying new onset autoantibodies in patients with COVID-19 (4), our identification of elevated anti-dsDNA IgG, which are associated with cardiovascular symptoms in systemic lupus (23), may provide a potential pathophysiological rationale for cardiovascular risk factors in severe COVID-19 (24). Indeed, two out of four class lll patients with vascular complications had increased levels of anti-dsDNA IgG in the CSF. Our observation of elevated anti-gut microbial IgA antibodies supports the evidence for gut barrier dysfunction in severe COVID-19 (25,26) that may necessitate containment of gut microbiota translocated to the circulation/CSF (27). In that regard, underlying conditions for microbiota dysbiosis, such as increased age, hypertension and diabetes have been observed in our class III cohort **(Table 1)**. Alternatively, trafficking of commensal-reactive regulatory B cells to sites of neuroinflammation as recently described (28,29) may underlie these findings. Our observations shed new light on mucosal barrier disruption as a modulator of the peripheral host immune response. While we cannot exclude that the differences in class III are due to pre-existing antibody profiles, the class-dependent increase in polyclonal antibody responses argues for a COVID-19-related pathophysiology and is corroborated by recent findings identifying new onset autoantibodies in patients with COVID-19 (4). In line with prior research (7,8), we observed a class-incremental cytokine storm in plasma, but less prominent in CSF. Intriguingly, class III patients displayed a unique CSF protein pattern highlighting BBB disruption, microglia regulation and neuronal tissue damage. TNFRSF12A displayed a high CSF-plasma correlation and predictive value for class III, rendering it a predictive biomarker for severe Neuro-COVID given its involvement in BBB disruption during CNS immune cell recruitment (30,31). Further, IL-8 (32), VEGFA (33) and EN-RAGE (34) promoted class III inherent BBB impairment, reinforcing ingress of polyreactive antibodies into the CSF.

Higher CSF protein, albumin, CSF/plasma ratio and IgM, IgG and IgA levels, and intrathecal detection of peripherally produced S-antibodies underscored BBB impairment in class III patients. Of note, S-antibody levels increase with a decreasing viral load (35), explaining our low S-antibody detection rate.

Exploiting microglia, neuronal markers and neuroimaging, we investigated consequences of COVID-19-induced BBB impairment on cerebral integrity. TNFRSF11B, a decoy receptor for RANKL, was the sole CSF discriminant between class III and ICs, leading to microglia overstimulation (36). Importantly, we detected concurring elevated RANKL CSF/plasma ratios in class III, which underscores the relevance of increased TNFRSF11B levels. Targeting TNFRSF11B, e.g. by RANKL mimics (37), could attenuate microglia activity in Neuro-COVID. Another relay of propagating the peripheral inflammation to the brain is represented by elevated MSR1 (38) followed by elevated CD200R1 in class III. Altogether, this suggests microglial activation and its consequences in severe Neuro-COVID.

Brain imaging revealed severe findings in class III patients, while class I/II patients lacked evidence of active neuroinflammation. Further, GMVs of olfactory pathway regions were decreased and correlated with CSF/blood albumin ratio, CSF leukocytes and protein in COVID-19 patients, accompanied by consistent CPVs. This result comes with the caveat that we included only 6 class III patients for CPV analysis (39).

Decreased GMVs in class II/III patients were associated with overexpression of the immune checkpoint protein PD-L1. Potentially, PD-L1 blockade would counteract this immune dysregulation and their associated structural brain changes (40).

Conversely, HGF, reported to mediate tissue-regenerative responses in COVID-19-induced lung damage (8), might serve as a counter-regulatory factor promoting neuroregeneration upon neuronal damage.

In contrast, lower plasma levels of neuroprotective GDF-8 (20) and BMP-4 (21), were associated with class II/III-related GMV loss, emphasizing the impact of COVID-19-induced soluble factors on distinct brain regions. Taken together, these GMV-associated CSF/plasma parameters may serve as targets to prevent long-term Neuro-COVID.

## Limitations

Although prospectively designed, we do not provide longitudinal follow-up data of assessed parameters. However, we provide a 10 months questionnaire-based follow-up confirming long-term neurological sequelae and higher mortality rates in class III.

Further, we recruited a relatively low number of class II patients, precluding us from characterizing this class to the same extent as class I/III.

Only unvaccinated patients were included since we recruited before the roll-out of COVID-19 vaccinations. Studies on the impact of vaccinations on reported findings might be of clinical relevance.

Finally, GMVs associations with distinctive biomarkers need cautious interpretation. While we detected significant associations between class I and class II/III patients, confounding factors such as age or co-existing morbidities were not considered.

## Conclusion

We provide a multiparametric framework of Neuro-COVID severity classifiers. Main determinants of severe Neuro-COVID are: (1) peripherally induced cytokine derangements, followed by (2) impaired BBB with ingressing polyreactive autoantibodies, (3) microglia reactivity and neuronal damage resulting in (4) potential GMV loss **(Figure S8)**. Collectively, these data identified several targets with the potential to prevent COVID-19-related neurological sequelae.

## Supporting information

Supplementary Methods, Figures and Tables description

## Data Availability

All data produced in the present work are contained in the manuscript

## Acknowledgments

We thank the patients and their relatives for their willingness to participate in this trial. We thank the medical and nursing staff of the University Hospital Basel Intensive Care Unit and COVID-19 cohort wards, as well as the Neuroradiology and Neurosurgery Departments for their huge efforts in making this trial possible. We thank Hedda Wardemann for providing us with the polyreactive ED-38 antibody clone and Emma Allen-Vercoe for providing us with the microbial template.

## Author contributions

Conceptualization: G.H., MM.E.

Methodology: G.H., MM.E., TA.M., L.K., E.P., A-K.P.

Clinical data and patient integration: G.H., MM.E., TA.M., J.R., N.E., C.E., I.J., E.K., H.P., M.S., A-K.P.

Sample handling and processing: MM.E., TA.M.

Software: W.D., S.H.

Formal analysis: W.D., S.H., JM.L., M-N.P., G.H., MM.E, TA.M., E.P., L.K., A-K.P.

Investigation: MM.E., TA.M., L.K., E.P.

Resources: N.E., J.R., M.S., M.K., I.J., H.P., M.S., J.O., J.K.

Writing - original draft: G.H., MM.E., TA.M., L.K., E.P., A-K.P.

Writing - review and editing: all authors

Supervision: G.H., A-K.P., M-N.P.

Project administration: G.H., A.T., M.M.

Funding acquisition: G.H., A.T., M.M.

