## Supplementary Methods, Figures and Tables description for "Severe Neuro-COVID is associated with peripheral immune signatures, autoimmunity and signs of neurodegeneration: a prospective cross-sectional study"

### Supplementary material

#### Supplementary methods

##### CSF and plasma sampling

Out of 40 patients 35 donated paired blood and CSF samples, whereas 5 participants donated only blood samples. Lumbar puncture and blood withdrawal were performed concomitantly, on an average latency period of 4 days after the first positive SARS-CoV-2 qRT-PCR test result. Lumbar puncture was performed under sterile conditions using a 20 gauge needle under local anesthesia on lumbar midline levels L4/5. Patients were monitored for positional headache or signs of CSF leakage for 24 h after puncture. Fresh CSF and EDTA-treated blood samples were processed into CSF supernatant and plasma. CSF samples were processed within 30 min post collection. After centrifugation at 1000 x *g* for 10 min, cell-free supernatant was removed, aliquoted and stored at -80°C. Whole blood was first centrifuged at 2000 x *g* for 10 min to separate plasma and blood cells. The isolated plasma was then centrifuged at 1000 x *g* for 10 min to remove residual blood cells, aliquoted and stored at -80°C. Retrospectively biobanked, age- and sex-matched paired CSF and plasma samples from patients with non-MS inflammatory neurologic disorders (n=25) and healthy patients (n=25) **(Table S1)** served as controls and were obtained from J.K. and J.O., Neurology Department, University Hospital Basel. The control groups are referred to as “HCs'' and “ICs''.

##### Antibody analysis

###### Quantification of total immunoglobulins and SARS-CoV-2 spike antibodies

Immunoglobulin (Ig) levels and anti-SARS-CoV-2 spike (S) protein IgG in plasma and CSF were quantified using nephelometric and ELISA assays and AI indices were calculated as part of the clinical routine diagnostic.

###### Anti-MOG and anti-NF155 antibody assays

Paired plasma and CSF supernatant samples from COVID-19 patients, HCs and ICs were examined for IgG reactivities against conformational human myelin oligodendrocyte glycoprotein (MOG)and neurofascin-155 (NF155) (41–44) using cell-based assays as previously described (41). In brief, stably transfected TE cells expressing full-length MOG, NF155 or the respective empty vector control were incubated with plasma (1:100) or CSF (1:5) and antibody binding was detected using secondary anti-human IgG-PE (Jackson). Humanized MOG- (h818C5) or NFSC-155-specific (A12/18.1) monoclonal antibodies were included as positive controls, respectively. Live cells were measured on a CytoFLEX flow cytometer and data analysis was performed in FlowJo (FlowJo 10.6.2, Becton Dickinson and Company). The ratio of the geometric mean channel fluorescence (MCF) of the transfected cell line divided by the MCF of the control cell line was calculated. The cut-off was set to 3 standard deviations above the mean of a HC cohort.

###### Commensal bacteria and polyreactivity ELISA

Human gut commensal bacteria, comprising 33 commensal bacteria strains (RePOOPulate) (29), double-stranded DNA (UltraPure Salmon Sperm DNA, Thermo Fisher) and bovine serum albumin (BSA, Sigma-Aldrich) were coated on a MaxiSorp ELISA plate (Nunc) in PBS in triplicates and incubated overnight at 4°C as recently reported (29). Plates were washed and blocked with 3% BSA in PBS for 2 hours at RT before incubation with plasma (1:100) or CSF (1:5) for 1 hour. After incubation with anti-human IgG or IgA horseradish peroxidase (Jackson ImmunoResearch) for 1 hour, the assay was developed with TMB peroxidase substrate (Seracare). A polyclonal polyreactive IgG antibody (ED38) was used as a positive assay control (45). Triplicates with a coefficient of variation (CV) greater than 15% were corrected for by excluding one value. Corrected duplicates with a CV above 15% were excluded from the analysis (n=4). Negative control signals (secondary antibodies only) were subtracted in a plate-specific manner.

##### Multiplexed secreted protein assays from CSF and plasma

A total of 192 analytes, including chemokines, soluble cell membrane proteins and cytokines, were measured in 85 paired plasma and CSF supernatant samples, consisting of COVID-19 patients (n=35), HCs (n=25) and patients with non-COVID-19 non-MS inflammatory neurological disorders (n=25). Additional unpaired plasma samples from COVID-19 patients (n=5) from whom CSF samples were not collected were also included. The Olink 96 target neurology (<https://www.olink.com/products-services/target/neurology-panel/>) and Olink 96 target inflammation (<https://www.olink.com/products-services/target/inflammation/>) panels were used. The measurements were performed by the Olink Analysis Service at Olink laboratories (SIAF, Davos, Switzerland). The assay used oligonucleotide-labeled antibody pairs allowing for pair-wise binding to target proteins. Briefly, when antibody pairs bound target antigens, corresponding oligonucleotides hybridized and were extended by polymerases and formed a unique barcode, allowing the quantification of protein analytes by high-throughput RT-PCR. Data are presented as normalized protein expression values, Olink Proteomics’ arbitrary unit on a log_2_ scale. Missing data were associated with a lower median expression. They were imputed as either half the molecule detection threshold or such that the sum of all imputed values for a molecule is 0.1 of the sum of the molecule's expressions, whichever was the smallest.

##### Analysis of multiplexed protein expression data

##### Software and code availability

The molecular data analysis was realized using python tools and libraries (Python Software Foundation, [http://www.python.org](http://www.python.org/)). Statistical computations relied on the numpy (46), pandas , scikit-learn, scipy and statsmodels libraries. Figures were generated using the graphviz, matplotlib and seaborn libraries. The code generated is available as a set of jupyter notebooks upon request to the corresponding author.

##### Handling of missing data/imputation

All values below the molecule-specific detection thresholds provided with the experimental results were treated as missing data which were imputed as either half the detection threshold or such that the sum of all imputed values on the column is 0.1 of the column values sum, whichever was the smallest.

##### Marginalization of age and sex

To account for discrepancies in the distribution of age and sex between the different groups, we used a linear model on the normalized protein expression (NPX) values, and marginalized individual observations for the median age and the female sex. Marginalization reports for each molecule are documented and available on request.

##### Single cytokine analysis

Across all measured molecules, only a minority followed the assumption of normality. Consequently, we relied on the Mann-Whitney-U test to detect significant differences in marginalized NPX values between our study cohorts. The test p-values were corrected using a BH-procedure to control the false discovery rate (FDR).

##### CSF/plasma ratio analysis

We investigated the relative concentration of each molecule between CSF and plasma (CSF/plasma index) to investigate whether they result from intrathecal or peripheral synthesis. Therefore, we first performed a median normalization of each molecule's NPX value. Significant differences in average ratios between groups were assessed using a Mann-Whitney-U test with a BH correction to control for the FDR.

##### Correlation analysis

The correlations between plasma and CSF measurements (between fluid correlations) were measured, for each COVID-19 class and control group, using the Kendall rank correlation coefficient (Kendall’s tau).

To correlate between CSF or plasma molecules and brain region volumes, we selected the brain regions (n=20, **Table S9)** and molecules **(Table S11,** *plasma* and *CSF*), which differed significantly between COVID-19 patients and controls. A Spearman-rank correlation test was performed to assess the association between regional brain volumes and marginalized protein expression levels in CSF and plasma. The BH procedure was used for FDR control.

The correlation between antibody levels and CSF/plasma proteins was performed using Spearman-rank correlation test, corrected with a BH procedure. The analysis was performed using the proteins' marginalized expression levels for the same proteins already selected for the regional brain volumes correlation analysis.

##### ROC-AUC analysis

We ranked molecules by the ROC-AUC score of their marginalized data to discriminate against either: class I versus (vs) III, class II vs III, class I+II vs III **(Table S7)**. Across all molecules, only a minority was normally distributed. Consequently, we relied on the nonparametric Mann-Whitney-U test to detect significant differences between each group. Individual p-values were corrected using the BH-procedure, controlling for an FDR of 0.05.

#### Brain imaging

Imaging studies were conducted on a 1.5 Tesla (T) MAGNETOM Siemens Avanto Fit and a 3T MAGNETOM Siemens Skyra Scanner. MRI sequences included 3D T1-weighted (T1w) +/- gadolinium, fluid-attenuated inversion recovery (FLAIR), diffusion-weighted imaging (DWI), susceptibility-weighted imaging (SWI) and T2-weighted (T2w) sequences to document signs of neuroinflammation. For standardization of MRI interpretation, an assessment protocol was created **(Table S2)**. Anatomical T1w magnetization prepared - rapid gradient echo (MPRAGE) pulse sequences were acquired for brain volumetric analysis. Cranial computerized tomography (CT) scans were assessed according to clinical standards. Two neuroradiologists (J.M.L. and M-N.P.) reviewed the images, blinded to clinical and laboratory patient data.

#### Brain volumetric analysis

##### Participants and imaging data acquisition

Among the 40 enrolled patients, 22 were selected based on the 3D high-resolution T1w anatomical image quality. To generate a bigger sample size, 13 additional patients who underwent brain MRI during their acute phase of COVID-19 were retrospectively added. These patients were not included in the main study cohort, undergoing CSF and plasma analysis. As a control cohort, 36 healthy, age- and sex-matched individuals were selected. This control group served only for the imaging analyses and was a different group than the HCs for the CSF and plasma comparison. The control groups’ demographic and clinical information can be found in **Table S12**. The 3D high-resolution T1w anatomical images were acquired using two MRI scanners (scanner 1: 1.5T Siemens Avanto Fit; scanner 2: 3T Siemens Skyra). An MPRAGE pulse sequence covering the whole brain was used in both MRI scanners with the following parameters. Scanner 1: 160 contiguous slices of 1 mm thickness in sagittal orientation; in-plane FOV=256 × 256 mm^2^, and matrix size 256 × 256 yielding an in-plane spatial resolution of 1 × 1 mm^2^ and voxel size of 1 × 1 × 1 mm^3^. The echo (TE), repetition (TR), and inversion (TI) times were set to TE/TR/TI=2.8 ms/2400 ms/900 ms with a flip angle FA=8°. Scanner 2: 160 contiguous slices of 1 mm thickness in sagittal orientation; in-plane FOV=256 × 240 mm^2^, and matrix size 256 × 240 yielding an in-plane spatial resolution of 1 × 1 mm^2^ and voxel size of 1 × 1 × 1 mm^3^. The echo, repetition, and inversion times were set to TE/TR/TI=2.98 ms/2300 ms/900 ms with a flip angle FA=9°. For the association analysis with the brain’s regional volume, we included routine diagnostic CSF parameters (leukocytes, lactate, protein, and blood albumin ratio). These variables were available for the main COVID-19 study cohort, which was under CSF and plasma evaluation.

##### Data pre-processing: brain global, regional gray matter and choroid plexus volume computation

The anatomical T1w images were automatically parcellated into 132 brain regions based on Neuromorphometrics atlas using the Neuromorphometrics toolbox. In short, the atlasing methodology consists of two main steps. First, each image is segmented into three different brain tissue classes (CSF, gray matter, and white matter) using the “Segment” (unified segmentation) tool in SPM12 (Statistical Parametric Mapping Toolbox), which includes registration to the MNI (Montreal Neurological Institute) space. Second, the probabilistic atlas of each of the anatomical structures is spatially registered with the extracted gray and white matter tissue maps using the “Shoot” tool in SPM12, based on a nonlinear advanced registration algorithm (47). Rules of probability are used to combine the previous images to obtain a probabilistic label map for each brain structure. At every gray matter voxel (in subject space), the probability of belonging to a specific anatomical structure is provided. From above, maximum probability label maps are calculated at all gray matter voxels (in subject space) which are labeled according to the structure of maximum probability. Finally, mean gray matter volume (GMV) values are calculated across voxels belonging to each structure label **(Table S13)**. The total intracranial volume (TIV) was computed as the sum of gray and white matter and cerebrospinal fluid volumes in cm^3^. Normalized GMV is defined as the ratio between gray matter volume and TIV.
To measure the choroid plexus volume (CPV) we segmented the choroid plexus of the lateral brain ventricles fully automatically on 3D T1-weighted MPRAGE sequences using a deep learning algorithm (Multi-Dimensional Gated Recurrent Units). To control for head size, we adjusted the statistical models for total intracranial volume (TIV), measured by SPM12.

#### Supplementary tables

##### **Table S1:** Characteristics of patients with a non-MS inflammatory neurologic disorder and HCs.

Demographics of HC patients and non-MS inflammatory neurologic disorder patients. For non-MS inflammatory neurologic disorder patients, specific neurological conditions are depicted.

##### **Table S2:** Brain imaging questionnaire.

For standardization of MRI interpretation, an assessment protocol was created. Cranial CT scans were assessed according to clinical standards.

##### **Table S3:** Detailed routine CSF parameters for each COVID-19 patient.

Detailed CSF parameters assessed during routine diagnostics are represented for each COVID-19 patient. Assessed parameters include glucose serum, lactate serum, CSF leukocytes, CSF erythrocytes, CSF lactate, CSF glucose, lactate and glucose ratio, protein total, CSF ferritin, Albumin CSF/plasma ratio, IgG index, IgG, IgA, IgM in the CSF, oligoclonal IgG bands in CSF vs serum and CSF-specific oligoclonal IgG bands.

##### **Table S4:** PCA components.

PC0 and PC1 components for both sample sources and merged exploratory proteomic panels (inflammatory and neurological proteins).

##### **Table S5:** Log_2_ scaled CSF/plasma ratio for each individual analyte and differences across study cohorts.

Differences in the CSF/plasma ratio for each protein and group.

##### **Table S6:** Nomenclature of relevant proteins in the study.

Naming of relevant study proteins according to the HUGO Gene Nomenclature Committee (HGNC, genenames.org).

##### **Table S7:** ROC-AUC analysis of individual proteins in different Neuro-COVID classes.

ROC-AUC analysis of individual CSF and plasma proteins and different group comparisons for prediction of class III development (class I versus III, class II versus III, class I and II versus III).

##### **Table S8:** Brain MRI and cranial CT results per Neuro-COVID class.

Detailed brain imaging analysis of structural brain imaging. Imaging analysis results are listed per Neuro-COVID class. CT scan results are described according to clinical standards.

##### **Table S9:** Smaller regional brain volumes in COVID-19 patients compared to the volumetric imaging control group.

##### Twenty brain regions were constituted with a lesser volume in the Neuro-COVID group compared to HC cases. Statistics: there were no significant differences after FDR correction.

##### Legend: std: standard deviation, uncorr: uncorrected, corr: corrected. Significant correlation values, p <0.05 (uncorrected), are represented in bold.

##### **Table S10:** Significant associations of regional brain volumes and clinical variables.

The significant associations between regional brain volume and clinical variables in the SARS-CoV-2 group. All correlation values were negative. For each region is shown the partial correlation value and their corrected p-value (FDR).

Legend: In bold, significant FDR p-values corrected (p <0.05). CSF: Cerebrospinal fluid; pval: p values corrected by FDR; Corr: correlation.

##### **Table S11:** Marginalized individual protein values and contrasts between different groups.

Marginalized values of 192 analytes, including chemokines, soluble cell membrane proteins and cytokines, were measured in 85 paired plasma and CSF supernatant samples and 5 additional plasma samples (35 paired samples of COVID-19 patients, 5 plasma samples of COVID-19 patients, 25 paired samples of non-MS inflammatory neurological disorder controls and HCs each). Individual NPX values and NPX values differences for each protein and group are represented.

##### **Table S12:** Main demographics and clinical variables of the COVID-19 imaging group and the volumetric imaging control group.

T-test for independence by group was used if the variable was normally distributed (Shapiro-Wilk, p <0.05). Non-parametric Mann-Whitney-U-test was applied whenever a variable for each group was not normally distributed, or there was no homogeneity of variances (Levene’s test); chi-square test for sex. Mean and standard deviation are shown for all variables except the categorical ones: sex, stroke, died in the hospital, Neuro-COVID state and MRI magnetic Field strength.

Legend: sd: standard deviation, CSF: Cerebrospinal fluid, TIV: Total Intracranial Volume, BPF: brain parenchymal fraction, na: not applicable. *****p <0.05.

##### **Table S13:** Regional brain volume values for the volumetric imaging COVID-19 group and control group.

Brain regional volume differences between SARS-CoV-2 patients and Control group. Legend: std: standard deviation, uncorr: uncorrected, corr: corrected. Significant correlations values, p<0.05 (uncorrected), are represented in bold.

#### Supplementary figures

##### **Figure S1:** Antibody reactivities against known CNS myelin antigens and total Ig and albumin values per Neuro-COVID class.

Box plot representations of anti-(non)-self reactivities in plasma, total Ig and albumin levels in plasma and CSF, and anti-myelin plasma reactivities. **A)** Anti-BSA, -dsDNA, anti-gut bacteria (RePOOPulate) IgG/IgA reactivities (OD450 nm) in plasma. **B)** Total CSF IgG/A/M and albumin levels (mg/L). Blue boxes indicate the clinical reference area.**C)** Plasma IgG reactivities against hMOG and NF155, plotted as geometric mean channel fluorescence (MCF) ratio. Dotted line indicates cut-off.

##### **Figure S2:** Heatmap of individual CSF and plasma analytes.

Z-score clustered heatmap visualization for each patient, group and sample source. **A)** Plasma analytes are represented on the X-axis. Each quadrant represents a single protein and individual participant. Participants and groups are represented on the Y-axis. Z-scores are color-coded with red colors *(high)*, white *(neutral)* and blue colors *(low).* **B)** CSF analytes are represented on the X-axis. Each quadrant represents a single protein and individual participant. Participants and groups are represented on the Y-axis. Z-scores are color-coded with red colors *(high)*, white *(neutral)* and blue colors *(low)*.

##### **Figure S3:** Low CSF/plasma soluble protein ratios are prevalent in Neuro-COVID patients indicating peripheral synthesis, whereas IC patients display predominant intrathecal changes.

**A)** Rose plots illustrating the log_2_-fold change of the CSF/plasma ratio for each cytokine in each group. A value of 0 corresponds to equality, a value of -1 refers to a 2 times less CSF concentration, a value of 1 means a 2 times less plasma concentration. For better visualization, the analytes were split into two separate rose plots. **B)** CSF/plasma ratios of each molecule were assessed to identify significant differences across groups. The ratio of 49 molecules significantly differed between groups after BH-procedure. These molecules are represented on the Y-axis on the right of the heatmaps. Ratio values are color-coded with red colors illustrating a higher ratio and blue colors illustrating a lower ratio. The 3 vertical differently colored lines on the Y-axis demonstrate specific ratio differences between *(from left to right)* class III vs ICs, class II vs healthy, and class I vs HCs. Heatmaps are separated into molecules with a *CSF/plasma ratio ≥0* *(upper heatmap)*, indicating intrathecal synthesis, and a *CSF/plasma ratio ≤0 (lower heatmap)*, indicating peripheral synthesis. The main ratio changes in Neuro-COVID patients take place in the plasma, whereas ICs display a stronger intrathecal immune response *(1)* compared to Neuro-COVID patients and *(2)* compared to immune reactions in their plasma.

##### **Figure S4:** Individual CSF and plasma analytes discriminating different groups.

Box plot representations of marginalized NPX of individual analytes significantly discriminating selected groups. **A)** Plasma, increasing NPX from *class I to III, and higher than in controls*: IL-6 (class III vs I: d=2.72, adj. p=0.007, class III vs infl. ctrl d=2.71, adj. p=0.001), IL-8 (class III vs I: d=1.4, adj. p=0.003, class III vs infl. ctrl d=1.8, adj. p=0.0002 ), HGF (class III vs I: d=1.36, adj. p=0.04, class III vs infl. ctrl d=1.76, adj. p=0.0007), VEGFA (class III vs I: d=0.6, adj. p=0.01, class III vs infl. ctrl d=0.8, adj. p=0.0005), EN-RAGE (class III vs I: d=2.86, adj. p=0.003, class III vs infl. ctrl d=3.34, adj. p=0.0002), TNFRSF12A (class III vs I: d=1.12, adj. p=0.006, class III vs infl. ctrl d=1.15, adj. p=0.002), PD-L1 (class III vs I: d=0.5, adj. p=0.04, class III vs infl. ctrl d=0.93, adj. p=0.002), CCL23 (class III vs infl. ctrl d=0.94, adj p = 0.01), EZR (class III vs infl. ctrl d=0.87, adj. p=0.002), TNFRSF11B (class III vs infl. ctrl d=0.34, adj. p=0.049). **B)** Plasma, decreasing NPX from *class I to III, and higher in controls than in COVID-19*: BMP-4, CLEC10A, CNTN5, GDF-8, NTRK2, GDNFRalpha, ROBO2. **C)** Plasma, Neuro-COVID class-independent, *higher NPX in COVID-19 than in controls:* 4E-BP1 (class III vs infl. ctrl d=1.19, adj p=0.007). **D)** Plasma, *higher NPX in COVID-19 than in controls and decreasing from class I to III:* HAGH (class III vs I: d=-0.98, adj. p=0.008). **E)** CSF, *increasing NPX from class I to III:* IL-8 (class III vs I: d=1.79, adj. p=0.012), MSR1 (class III vs I: d=1.01, adj p=0.016), 4E-BP1 (class III vs I: d=1.16, adj p=0.02), CD200R1 (class III vs I: d=0.50, adj p=0.04), TNFRSF12A (class III vs I: d=1.16, adj p=0.008), EZR (class III vs I: d=0.76, adj p=0.01). **H)** CSF, *increasing NPX from class I to III, and higher in class III than in ICs:* TNFRSF11B (class III vs I: d=0.8, adj. p=0.04, class III vs infl. ctrl d=0.98, adj. p=0.02).

Statistics: statistical significance was calculated using the Mann-Whitney-U test (adj. p: *<0.05, **<0.01, ***<0.001).

Legend: NPX: normalized protein expression, d: difference, adj. p: adjusted p-value, NS: not significant.

##### **Figure S5:** Map of brain regions significantly correlating with CSF parameters.

Map of the 16 brain regions with significant correlation values of GMV and clinical variables in the Neuro-COVID group after multiple comparison correction (FDR). These regions are represented in different colors on a T1-weighted template (in radiological convention).

Legend: L: left, R: right.

##### **Figure S6:** Elevated GDF-8 plasma levels in class I are associated with preserved regional brain volumes, whereas higher HGF plasma levels in class III are associated with decreased regional brain volumes.

**A)** Correlation plots demonstrating the correlation of GDF-8 and different regional brain volumes in Neuro-COVID patients. The Y-axis represents the regional brain volume values. The X-axis represents the marginalized NPX of the respective protein. None of the adjusted p-values was significant after BH-procedure. **B)** Boxplot representations of marginalized NPX of class I compared to merged class II+II plasma GDF-8 (Wilcoxon, p=0.025)*.* **C)** Correlation plots demonstrating the correlation of HGF and different regional brain volumes in Neuro-COVID patients. The Y-axis represents the regional brain volume values. The X-axis represents the marginalized NPX of the respective protein. None of the adjusted p-values was significant after BH-procedure. **B)** Boxplot representations of marginalized NPX of class I compared to merged class II+II plasma HGF (Wilcoxon, p=0.06).

Legend: RightFO: right frontal operculum, LeftPrG: left precentral gyrus, RightOpIFG: right opercular part of the inferior frontal gyrus, RightInfLatVent: right inferior lateral ventricle, LeftPP: left planum polare, RightPIns: right posterior insula, RightCO: right central operculum.

##### **Figure S7:** Correlation analysis of brain regions and proteins with most significant associations.

**A)** Boxplot representations of marginalized NPX individual plasma analytes associated with *decreased regional brain volumes:* PD-L1, HGF, CX3CL1, IL-15RA, EN-RAGE; and plasma analytes associated with *protective effects on regional brain volumes:* GDF-8, BMP-4, NTRK2. Statistics: none of the p-values are significant after BH-procedure. **B)** Boxplot representations of marginalized NPX individual CSF analytes associated with *decreased regional brain volumes:* EZR, IL-8, 4E-BP1; and CSF analytes associated with *protective effects on regional brain volumes:* EZR, NTRK2, ROBO2, RGMB, CD200. Statistics: none of the p-values are significant after BH-procedure.

Legend: RightFO: right frontal operculum, LeftPrG: left precentral gyrus, RightOpIFG: right opercular part of the inferior frontal gyrus, RightInfLatVent: right inferior lateral ventricle, LeftPP: left planum polare, RightPIns: right posterior insula, RightCO: right central operculum, R: correlation coefficient, p: p-value.

##### **Figure S8:** Overview of proposed pathomechanisms leading to Neuro-COVID.

The proposed main determinants of severe Neuro-COVID are: (1) peripherally induced cytokine derangements, followed by (2) impaired BBB with ingressing polyreactive autoantibodies, resulting in (3) microglia reactivity and neuronal damage. Created with BioRender.com.
