## Supplementary material for "Severe Neuro-COVID is associated with peripheral immune signatures, autoimmunity and signs of neurodegeneration: a prospective cross-sectional study": Table S1

| **Healthy control cases** (n=25) | |
| --- | --- |
| **Age, years, mean (SD)** | 52 (18) |
| Range, years | 23-75 |
| **Sex** |  |
| Female, n (%) | 12 (48%) |
| Male, n (%) | 13 (52%) |
| **Inflammatory control cases** (n=25) | |
| **Age, years, mean (SD)** | 54 (19) |
| Range, years | 20-82 |
| **Sex** |  |
| Female, n (%) | 12 (48%) |
| Male, n (%) | 13 /52%) |
| **Neurologic disorder** |  |
| Postherpetic neuralgia, n (%) | 1 (4%) |
| Herpetic meningitis, n (%) | 1 (4%) |
| Herpetic encephalitis, n (%) | 1 (4%) |
| Herpetic meningoencephalitis, n (%) | 1 (4%) |
| VZV meningomyeloradiculitis, n (%) | 1 (4%) |
| Disseminated herpes zoster with CNS affection, n (%) | 3 (12%) |
| Viral meningitis (not specified), n (%) | 2 (8%) |
| Viral meningoencephalitis (not specified), n (%) | 1 (4%) |
| Relapsing meningitis of unclear etiology, n (%) | 1 (4%) |
| Eosinophilic encephalitis, n (%) | 3 (12%) |
| Cranial neuropathy, n (%) | 1 (4%) |
| Tuberculous meningoencephalitis, n (%) | 3 (12%) |
| Neuroborreliosis, n (%) | 3 (12%) |
| Red nucleus lesion of unclear etiology, n (%) | 1 (4%) |
| Neurosarcoidosis, n (%) | 1 (4%) |
| Susac’s syndrome, n (%) | 1 (4%) |
| Autoimmune encephalitis, n (%) | 1 (4%) |
| Rasmussen encephalitis, n (%) | 1 (4%) |
| SD: standard deviation  VZV: varicella-zoster virus  CNS: central nervous system | |
