## Supplementary material for "Severe Neuro-COVID is associated with peripheral immune signatures, autoimmunity and signs of neurodegeneration: a prospective cross-sectional study": Table S2

| **Study ID** |  |
| --- | --- |
| **MRI field force** |  |
| **DWI lesions** |  |
| Gray matter | Yes/no |
| If yes – quantification |  |
| White matte | Yes/no |
| If yes – quantification |  |
| Temporal lobe | Yes/no |
| If yes – quantification |  |
| Periventricular region | Yes/no |
| If yes – quantification |  |
| Basal ganglia | Yes/no |
| If yes – quantification |  |
| Corpus callosum | Yes/no |
| If yes – quantification |  |
| Brain stem | Yes/no |
| If yes – quantification |  |
| Cerebellum/cerebellar peduncles | Yes/no |
| If yes – quantification |  |
| DWI lesions | Homogeneous/heterogeneous |
| Majority of DWI lesions | Hypointense/isointense/hyperintense in ADC |
| Additional remarks |  |
| **FLAIR/T2w imaging lesions** |  |
| White matter | Yes/no |
| If yes - quantification |  |
| Semioval center | Yes/no |
| If yes – quantification |  |
| Temporal lobe | Yes/no |
| If yes – quantification |  |
| Mesiotemporal region | Yes/no |
| If yes – quantification |  |
| Periventricular region | Yes/no |
| If yes – quantification |  |
| Basal ganglia | Yes/no |
| If yes – quantification |  |
| Corpus callosum | Yes/no |
| If yes – quantification |  |
| Brain stem | Yes/no |
| If yes – quantification |  |
| Cerebellum/cerebellar peduncles | Yes/no |
| Additional remarks |  |
| **SWI imaging lesions** |  |
| Periventricular region | Yes/no |
| If yes – quantification |  |
| Basal ganglia | Yes/no |
| If yes – quantification |  |
| Thalamus | Yes/no |
| If yes - quantification |  |
| Internal capsule | Yes/no |
| If yes – quantification |  |
| External capsule | Yes/no |
| If yes – quantification |  |
| Corpus callosum | Yes/no |
| If yes – quantification |  |
| Brain stem | Yes/no |
| If yes – quantification |  |
| Cerebellum/cerebellar peduncles | Yes/no |
| If yes – quantification |  |
| Frontal lobe | Yes/no |
| If yes – quantification |  |
| Temporal lobe | Yes/no |
| If yes – quantification |  |
| Parietal lobe | Yes/no |
| If yes – quantification |  |
| Occipital lobe | Yes/no |
| If yes – quantification |  |
| Insula | Yes/no |
| If yes – quantification |  |
| Majority of SWI lesions | Microhemorrhagic (< 10mm), macrohemorrhagic foci |
| Laterality | Unilateral/bilateral |
| Other remarks |  |
| **Gadolinium enhanced T1w imaging** |  |
| Thrombosis signal | Yes/no |
| Meningeal/leptomeningeal enhancement | Yes/no |
| If yes | Focal/diffuse, supratentorial/infratentorial |
| Parenchymal enhancement | Yes/no |
| If yes - quantification |  |
| **Olfactory bulbus imaging** |  |
| Signal alterations on FLAIR/T2w imaging | Yes/no |
| Signal alterations on T1w imaging | Yes/no |
| Signal alterations on T1w gadolinium enhanced imaging | Yes/no |
| MRI: magnetic resonance imaging  DWI: diffusion weighted imaging  ADC: apparent diffusion coefficient  FLAIR: fluid attenuated inversion recovery  T2w: T2-weighted  SWI: susceptibility-weighted imaging  T1w: T1-weighted | |
