## Supplementary material for "Severe Neuro-COVID is associated with peripheral immune signatures, autoimmunity and signs of neurodegeneration: a prospective cross-sectional study": Table S9

| **Structures** | **Control (mean)** | **Control (std)** | **Patient (mean)** | **Patient (std)** | **t stat** | **pval uncor** | **pval**  **corr** |
| --- | --- | --- | --- | --- | --- | --- | --- |
| **Right Hippocampus** | 3.321 | 0.388 | 3.360 | 0.349 | -2.227 | **0.035** | 0.297 |
| **Right Inf Lat Vent** | 0.697 | 0.302 | 0.923 | 0.754 | -2.132 | **0.043** | 0.297 |
| **Right Central Operculum** | 4.005 | 0.430 | 3.693 | 0.471 | 2.137 | **0.042** | 0.297 |
| **Right Frontal Operculum** | 2.027 | 0.264 | 1.843 | 0.260 | 2.288 | **0.031** | 0.297 |
| **Right Medial Frontal Cortex** | 1.889 | 0.234 | 1.780 | 0.290 | 2.222 | **0.036** | 0.297 |
| **Right Middle Frontal Gyrus** | 18.939 | 2.250 | 18.425 | 2.590 | 2.062 | **0.049** | 0.315 |
| **Left Postcentral Gyrus Medial Segment** | 1.184 | 0.180 | 1.102 | 0.159 | 2.313 | **0.029** | 0.297 |
| **Right Precentral Gyrus Medial Segment** | 2.667 | 0.355 | 2.510 | 0.347 | 2.213 | **0.036** | 0.297 |
| **Left Precentral Gyrus Medial Segment** | 2.714 | 0.371 | 2.524 | 0.375 | 3.237 | **0.003** | 0.190 |
| **Right Superior Frontal Gyrus Medial Segment** | 8.277 | 1.092 | 7.843 | 1.165 | 2.659 | **0.013** | 0.275 |
| **Left Superior Frontal Gyrus Medial Segment** | 7.376 | 0.873 | 7.006 | 0.980 | 2.246 | **0.034** | 0.297 |
| **Right Posterior Insula** | 2.371 | 0.292 | 2.164 | 0.285 | 2.362 | **0.026** | 0.297 |
| **Right Postcentral Gyrus** | 10.984 | 1.546 | 10.299 | 1.287 | 2.398 | **0.024** | 0.297 |
| **Right Planum Polare** | 2.101 | 0.238 | 1.946 | 0.228 | 2.331 | **0.028** | 0.297 |
| **Left Planum Polare** | 2.327 | 0.244 | 2.178 | 0.259 | 2.150 | **0.041** | 0.297 |
| **Right Precentral Gyrus** | 12.895 | 1.575 | 12.222 | 1.515 | 2.617 | **0.015** | 0.275 |
| **Left Precentral Gyrus** | 13.296 | 1.589 | 12.590 | 1.454 | 3.238 | **0.003** | 0.190 |
| **Right Supplementary Motor Cortex** | 5.461 | 0.713 | 5.243 | 0.712 | 2.698 | **0.012** | 0.275 |
| **Left Supplementary Motor Cortex** | 5.569 | 0.733 | 5.243 | 0.806 | 2.914 | **0.007** | 0.228 |
