## Supplementary material for "Severe Neuro-COVID is associated with peripheral immune signatures, autoimmunity and signs of neurodegeneration: a prospective cross-sectional study": Table S10

| Structures | CSF Leukocytes Corr | pval | CSF Protein Corr | pval | CSF Blood Albumin Ratio Corr | pval |
| --- | --- | --- | --- | --- | --- | --- |
| Left Accumbens Area | -0.821 | **0.029** | -0.709 | 0.054 | -0.767 | 0.059 |
| Left Amygdala | -0.687 | 0.081 | -0.744 | **0.049** | -0.740 | 0.078 |
| Left Basal Forebrain | -0.749 | 0.050 | -0.664 | 0.067 | -0.657 | 0.111 |
| Left Fusiform Gyrus | -0.721 | 0.060 | -0.788 | **0.036** | -0.750 | 0.073 |
| Left Inferior Temporal Gyrus | -0.761 | 0.050 | -0.599 | 0.104 | -0.704 | 0.082 |
| Left Medial Orbital Gyrus | -0.748 | 0.050 | -0.601 | 0.104 | -0.687 | 0.089 |
| Left Posterior Cingulate Gyrus | -0.746 | 0.050 | -0.593 | 0.105 | -0.528 | 0.266 |
| Optic Chiasm | -0.639 | 0.123 | -0.758 | **0.044** | -0.829 | **0.028** |
| Right Anterior Orbital Gyrus | -0.805 | **0.029** | -0.763 | **0.044** | -0.806 | **0.040** |
| Right Amygdala | -0.727 | 0.058 | -0.832 | **0.015** | -0.838 | **0.028** |
| Right Angular Gyrus | -0.432 | 0.329 | -0.688 | 0.062 | -0.837 | **0.028** |
| Right Entorhinal Area | -0.849 | **0.029** | -0.866 | **0.008** | -0.899 | **0.009** |
| Right Lateral Orbital Gyrus | -0.816 | **0.029** | -0.632 | 0.078 | -0.639 | 0.118 |
| Right Posterior Orbital Gyrus | -0.795 | **0.031** | -0.714 | 0.054 | -0.793 | **0.046** |
| Right Putamen | -0.588 | 0.157 | -0.731 | 0.049 | -0.786 | **0.046** |
| Right Superior Frontal Gyrus | -0.571 | 0.172 | -0.733 | 0.049 | -0.705 | 0.082 |
