## Supplementary material for "Severe Neuro-COVID is associated with peripheral immune signatures, autoimmunity and signs of neurodegeneration: a prospective cross-sectional study": Table S12

|  | COVID-19 | Controls |
| --- | --- | --- |
| Numbers of subjects | 35 | 36 |
| Age | 51.9, sd (19.7) | 54.03, sd (23.7) |
| Sex, Male/Female | M/F (14/21) | (13/23) |
| stroke | 2 | na |
| CSF Leukocytes | 17 (4.29, sd 3.72) | na |
| CSF Lactate | 18 (1.89, sd 0.50) | na |
| CSF Protein | n=18 (346.11, sd 180.83) | na |
| CSF Blood Albumin Ratio | n=15 (6.50, sd 4.60) | na |
| CSF Glucose | n=18 (4.49, sd 1.30) | na |
| Plasma TRANCE | n=20 (2.74, sd 0.93) | na |
| Plasma EN-RAGE | n=20 (3.30, sd 1.37) | na |
| CSF OPG | n=18 (9.55, sd 0.68) | na |
| CSF TRANCE | n=18 (-0.31, sd 0.29) | na |
| CSF EN-RAGE | n=18 (0.43, sd: 0.74) | na |
| Died in Hospital | 1 | na |
| Neurocovid state | n=1 (13), 2 (6), 3 (2) | na |
| Weight [kg] | 69.88 (sd 11.95) | 71.53(sd 16.30) |
| Height [meters] | 1.69 (sd 0.09) | 1.69 (sd 0.11) |
| MRI magnetic Field strength | 1.5 T (30)  3 T (5) | 1.5 T (11)  3 T (25) |
| TIV [cm^3^] | 1431(sd 141,96) | 1497(sd 156.293) |
| Global gray matter [cm^3^] | 618.65 (sd 98.86) | 638.61 (sd 95.38) |
| Global White matter [cm^3^]* | 449.28 (sd 63.76) | 451.09 (sd 62.63) |
| CSF matter [cm^3^] | 373.04 (sd 137.65) | 408.01 (sd 147.46) |
| BPF | 0.74 (sd 0.08) | 0.73 (sd 0.08) |
| Parenchyma Volume [cm^3^] | 1067.94 (sd 138.22) | 1089.70 (sd 137.40) |
