## Supplementary material for "Severe Neuro-COVID is associated with peripheral immune signatures, autoimmunity and signs of neurodegeneration: a prospective cross-sectional study": Table S13

| **Structures** | **Control (mean)** | **Control (std)** | **Patient (mean)** | **Patient (std)** | **t stat** | **pval uncor** | **pval**  **corr** |
| --- | --- | --- | --- | --- | --- | --- | --- |
| **Right Accumbens Area** | 0.399 | 0.056 | 0.377 | 0.064 | 1.355 | 0.159 | 0.397 |
| **Left Accumbens Area** | 0.434 | 0.063 | 0.414 | 0.064 | 1.208 | 0.191 | 0.397 |
| **Right Amygdala** | 0.864 | 0.124 | 0.857 | 0.106 | -1.039 | 0.231 | 0.397 |
| **Left Amygdala** | 0.874 | 0.103 | 0.858 | 0.112 | -0.667 | 0.317 | 0.397 |
| **Brain Stem** | 18.131 | 2.146 | 17.696 | 1.724 | 0.460 | 0.357 | 0.397 |
| **Right Caudate** | 3.122 | 0.427 | 2.883 | 0.484 | 1.572 | 0.116 | 0.340 |
| **Left Caudate** | 2.962 | 0.451 | 2.734 | 0.480 | 1.375 | 0.154 | 0.397 |
| **Right Cerebellum Exterior** | 47.606 | 5.393 | 45.845 | 5.015 | 0.670 | 0.317 | 0.397 |
| **Left Cerebellum Exterior** | 47.333 | 5.407 | 45.774 | 5.532 | 0.732 | 0.303 | 0.397 |
| **Right Cerebellum White Matter** | 14.661 | 2.021 | 14.579 | 1.819 | 0.477 | 0.354 | 0.397 |
| **Left Cerebellum White Matter** | 14.991 | 2.070 | 14.650 | 1.888 | 0.841 | 0.278 | 0.397 |
| **Right Cerebral White Matter** | 220.382 | 28.536 | 218.779 | 30.965 | 1.199 | 0.193 | 0.397 |
| **Left Cerebral White Matter** | 217.331 | 28.198 | 215.791 | 27.589 | 0.994 | 0.242 | 0.397 |
| **CSF** | 1.474 | 0.602 | 1.335 | 0.523 | -0.865 | 0.272 | 0.397 |
| **Right Hippocampus** | 3.321 | 0.388 | 3.360 | 0.349 | -2.227 | **0.035** | 0.297 |
| **Left Hippocampus** | 3.134 | 0.308 | 3.142 | 0.303 | -1.672 | 0.099 | 0.327 |
| **Right Inf Lat Vent** | 0.697 | 0.302 | 0.923 | 0.754 | -2.132 | **0.043** | 0.297 |
| **Left Inf Lat Vent** | 0.651 | 0.305 | 0.709 | 0.467 | -1.245 | 0.183 | 0.397 |
| **Right Lateral Ventricle** | 16.625 | 11.184 | 13.942 | 9.038 | -0.489 | 0.352 | 0.397 |
| **Left Lateral Ventricle** | 18.341 | 11.564 | 16.196 | 11.378 | -0.671 | 0.316 | 0.397 |
| **Right Pallidum** | 1.575 | 0.182 | 1.537 | 0.177 | 0.602 | 0.331 | 0.397 |
| **Left Pallidum** | 1.576 | 0.188 | 1.535 | 0.167 | 0.339 | 0.375 | 0.397 |
| **Right Putamen** | 3.926 | 0.510 | 3.721 | 0.502 | 1.973 | 0.058 | 0.315 |
| **Left Putamen** | 3.958 | 0.515 | 3.768 | 0.517 | 1.296 | 0.171 | 0.397 |
| **Right Thalamus Proper** | 6.982 | 0.797 | 6.920 | 0.769 | -0.180 | 0.391 | 0.397 |
| **Left Thalamus Proper** | 7.320 | 0.832 | 7.223 | 0.819 | -0.331 | 0.376 | 0.397 |
| **Right Ventral DC** | 4.612 | 0.442 | 4.561 | 0.411 | 0.329 | 0.376 | 0.397 |
| **Left Ventral DC** | 4.841 | 0.467 | 4.777 | 0.441 | 0.004 | 0.397 | 0.397 |
| **Optic Chiasm** | 0.077 | 0.007 | 0.075 | 0.009 | -0.124 | 0.394 | 0.397 |
| **Cerebellar Vermal Lobules I_V** | 4.720 | 0.546 | 4.548 | 0.469 | 0.784 | 0.291 | 0.397 |
| **Cerebellar Vermal Lobules VI_VII** | 2.231 | 0.210 | 2.157 | 0.223 | 0.828 | 0.281 | 0.397 |
| **Cerebellar Vermal Lobules VIII_X** | 2.673 | 0.319 | 2.608 | 0.290 | 0.453 | 0.358 | 0.397 |
| **Left Basal Forebrain** | 0.390 | 0.042 | 0.378 | 0.043 | 0.941 | 0.254 | 0.397 |
| **Right Basal Forebrain** | 0.400 | 0.044 | 0.380 | 0.044 | 1.632 | 0.106 | 0.340 |
| **Right Anterior Cingulate Gyrus** | 4.178 | 0.466 | 4.045 | 0.527 | 0.831 | 0.280 | 0.397 |
| **Left Anterior Cingulate Gyrus** | 4.838 | 0.585 | 4.622 | 0.654 | 1.423 | 0.144 | 0.388 |
| **Right Anterior Insula** | 4.146 | 0.470 | 3.824 | 0.432 | 1.893 | 0.068 | 0.315 |
| **Left Anterior Insula** | 4.335 | 0.421 | 4.058 | 0.487 | 1.087 | 0.219 | 0.397 |
| **Right Anterior Orbital Gyrus** | 1.859 | 0.259 | 1.815 | 0.299 | 0.972 | 0.247 | 0.397 |
| **LeftA Anterior Orbital Gyrus** | 1.643 | 0.237 | 1.609 | 0.231 | 0.860 | 0.273 | 0.397 |
| **Right Angular Gyrus** | 10.208 | 1.363 | 10.111 | 1.304 | -1.103 | 0.216 | 0.397 |
| **Left Angular Gyrus** | 9.164 | 1.180 | 9.136 | 1.095 | -0.251 | 0.385 | 0.397 |
| **Right Calcarine Cortex** | 3.392 | 0.443 | 3.267 | 0.458 | -0.516 | 0.347 | 0.397 |
| **Left Calcarine Cortex** | 3.266 | 0.438 | 3.149 | 0.487 | -0.795 | 0.289 | 0.397 |
| **Right Central Operculum** | 4.005 | 0.430 | 3.693 | 0.471 | 2.137 | **0.042** | 0.297 |
| **Left Central Operculum** | 4.141 | 0.436 | 3.860 | 0.527 | 1.813 | 0.078 | 0.315 |
| **Right Cuneus** | 4.796 | 0.560 | 4.727 | 0.626 | -0.410 | 0.365 | 0.397 |
| **Left Cuneus** | 4.646 | 0.581 | 4.409 | 0.670 | 0.436 | 0.361 | 0.397 |
| **Right Entorhinal Area** | 1.712 | 0.253 | 1.672 | 0.225 | -0.296 | 0.380 | 0.397 |
| **Left Entorhinal Area** | 1.685 | 0.196 | 1.639 | 0.218 | -0.109 | 0.395 | 0.397 |
| **Right Frontal Operculum** | 2.027 | 0.264 | 1.843 | 0.260 | 2.288 | **0.031** | 0.297 |
| **Left Frontal Operculum** | 2.037 | 0.228 | 1.933 | 0.287 | 1.277 | 0.175 | 0.397 |
| **Right Frontal Pole** | 3.996 | 0.452 | 3.844 | 0.465 | 1.542 | 0.122 | 0.340 |
| **Left Frontal Pole** | 3.723 | 0.423 | 3.618 | 0.446 | 0.850 | 0.276 | 0.397 |
| **Right Fusiform Gyrus** | 7.609 | 0.979 | 7.408 | 0.869 | -0.049 | 0.397 | 0.397 |
| **Left Fusiform Gyrus** | 7.422 | 0.852 | 7.173 | 0.881 | 0.765 | 0.296 | 0.397 |
| **Right Gyrus Rectus** | 2.152 | 0.253 | 2.115 | 0.334 | 0.965 | 0.249 | 0.397 |
| **Left Gyrus Rectus** | 2.146 | 0.240 | 2.148 | 0.315 | 0.336 | 0.375 | 0.397 |
| **Right Inferior Occipital Gyrus** | 6.444 | 0.878 | 6.419 | 0.897 | -1.685 | 0.097 | 0.327 |
| **Left Inferior Occipital Gyrus** | 6.345 | 0.938 | 6.164 | 0.785 | -0.008 | 0.397 | 0.397 |
| **Right Inferior Temporal Gyrus** | 11.568 | 1.516 | 11.219 | 1.379 | 0.857 | 0.274 | 0.397 |
| **Left Inferior Temporal Gyrus** | 11.352 | 1.317 | 10.902 | 1.290 | 1.816 | 0.078 | 0.315 |
| **Right Lingual Gyrus** | 8.326 | 0.897 | 7.871 | 1.004 | 0.577 | 0.336 | 0.397 |
| **Left Lingual Gyrus** | 7.661 | 0.833 | 7.301 | 0.969 | -0.056 | 0.397 | 0.397 |
| **Right Lateral Orbital Gyrus** | 2.371 | 0.299 | 2.273 | 0.320 | 1.531 | 0.124 | 0.340 |
| **Left Lateral Orbital Gyrus** | 2.526 | 0.314 | 2.454 | 0.331 | 0.633 | 0.324 | 0.397 |
| **Right Middle Cingulate Gyrus** | 4.683 | 0.581 | 4.703 | 0.829 | -0.071 | 0.396 | 0.397 |
| **Left Middle Cingulate Gyrus** | 4.811 | 0.594 | 4.832 | 0.791 | -0.024 | 0.397 | 0.397 |
| **Right Medial Frontal Cortex** | 1.889 | 0.234 | 1.780 | 0.290 | 2.222 | **0.036** | 0.297 |
| **Left Medial Frontal Cortex** | 1.906 | 0.237 | 1.838 | 0.288 | 1.202 | 0.192 | 0.397 |
| **Right Middle Frontal Gyrus** | 18.939 | 2.250 | 18.425 | 2.590 | 2.062 | **0.049** | 0.315 |
| **Left Middle Frontal Gyrus** | 19.269 | 2.169 | 18.924 | 2.490 | 1.546 | 0.121 | 0.340 |
| **Right Middle Occipital Gyrus** | 5.009 | 0.632 | 4.976 | 0.720 | -1.999 | 0.056 | 0.315 |
| **Left Middle Occipital Gyrus** | 6.038 | 0.895 | 6.035 | 0.806 | -1.728 | 0.090 | 0.327 |
| **Right Media lOrbital Gyrus** | 3.951 | 0.458 | 3.856 | 0.562 | 1.291 | 0.172 | 0.397 |
| **Left Medial Orbital Gyrus** | 4.098 | 0.473 | 4.053 | 0.529 | 0.741 | 0.301 | 0.397 |
| **Right Postcentral Gyrus Medial Segment** | 1.139 | 0.148 | 1.059 | 0.153 | 1.940 | 0.062 | 0.315 |
| **Left Postcentral Gyrus Medial Segment** | 1.184 | 0.180 | 1.102 | 0.159 | 2.313 | **0.029** | 0.297 |
| **Right Precentral Gyrus Medial Segment** | 2.667 | 0.355 | 2.510 | 0.347 | 2.213 | **0.036** | 0.297 |
| **Left Precentral Gyrus Medial Segment** | 2.714 | 0.371 | 2.524 | 0.375 | 3.237 | **0.003** | 0.190 |
| **Right Superior Frontal Gyrus Medial Segment** | 8.277 | 1.092 | 7.843 | 1.165 | 2.659 | **0.013** | 0.275 |
| **Left Superior Frontal Gyrus Medial Segment** | 7.376 | 0.873 | 7.006 | 0.980 | 2.246 | **0.034** | 0.297 |
| **Right Middle Temporal Gyrus** | 14.124 | 1.811 | 13.701 | 1.794 | 0.515 | 0.347 | 0.397 |
| **Left Middle Temporal Gyrus** | 13.853 | 1.849 | 13.321 | 1.564 | 1.719 | 0.092 | 0.327 |
| **Right Occipital Pole** | 2.665 | 0.410 | 2.664 | 0.508 | -1.773 | 0.084 | 0.315 |
| **Left Occipital Pole** | 3.137 | 0.437 | 3.102 | 0.451 | -1.789 | 0.081 | 0.315 |
| **Right Occipital Fusiform Gyrus** | 4.355 | 0.493 | 4.152 | 0.593 | 0.297 | 0.380 | 0.397 |
| **Left Occipital Fusiform Gyrus** | 4.380 | 0.523 | 4.182 | 0.582 | 0.694 | 0.311 | 0.397 |
| **Right Opercular Part Of The Inferior Frontal Gyrus** | 3.578 | 0.480 | 3.238 | 0.450 | 3.023 | **0.005** | 0.228 |
| **Left Opercular Part Of The Inferior FrontalGyrus** | 3.416 | 0.393 | 3.314 | 0.424 | 1.057 | 0.226 | 0.397 |
| **Right Orbital Part Of The Inferior Frontal Gyrus** | 1.612 | 0.190 | 1.531 | 0.217 | 1.677 | 0.098 | 0.327 |
| **Left Orbital Part Of The Inferior Frontal Gyrus** | 1.602 | 0.180 | 1.549 | 0.238 | 0.754 | 0.298 | 0.397 |
| **Right Posterior Cingulate Gyrus** | 4.056 | 0.466 | 4.034 | 0.534 | -0.735 | 0.302 | 0.397 |
| **Left Posterior Cingulate Gyrus** | 4.433 | 0.488 | 4.400 | 0.560 | -0.838 | 0.279 | 0.397 |
| **Right Precuneus** | 10.839 | 1.148 | 10.586 | 1.410 | 0.164 | 0.392 | 0.397 |
| **Left Precuneus** | 10.794 | 1.158 | 10.622 | 1.457 | -0.151 | 0.393 | 0.397 |
| **Right Parahippocampal Gyrus** | 2.974 | 0.329 | 2.933 | 0.323 | -0.772 | 0.294 | 0.397 |
| **Left Parahippocampal Gyrus** | 3.204 | 0.293 | 3.163 | 0.331 | -0.573 | 0.337 | 0.397 |
| **Right Posterior Insula** | 2.371 | 0.292 | 2.164 | 0.285 | 2.362 | **0.026** | 0.297 |
| **Left Posterior Insula** | 2.332 | 0.242 | 2.175 | 0.280 | 1.266 | 0.178 | 0.397 |
| **Right Parietal Operculum** | 2.256 | 0.290 | 2.048 | 0.354 | 1.820 | 0.077 | 0.315 |
| **Left Parietal Operculum** | 2.475 | 0.333 | 2.276 | 0.368 | 1.410 | 0.147 | 0.388 |
| **Right Postcentral Gyrus** | 10.984 | 1.546 | 10.299 | 1.287 | 2.398 | **0.024** | 0.297 |
| **Left Postcentral Gyrus** | 12.119 | 1.319 | 11.738 | 1.448 | 0.569 | 0.337 | 0.397 |
| **Right Posterior Orbital Gyrus** | 2.393 | 0.291 | 2.253 | 0.279 | 1.783 | 0.082 | 0.315 |
| **Left Posterior Orbital Gyrus** | 2.556 | 0.319 | 2.464 | 0.291 | 1.575 | 0.115 | 0.340 |
| **Right Planum Polare** | 2.101 | 0.238 | 1.946 | 0.228 | 2.331 | **0.028** | 0.297 |
| **Left Planum Polare** | 2.327 | 0.244 | 2.178 | 0.259 | 2.150 | **0.041** | 0.297 |
| **Right Precentral Gyrus** | 12.895 | 1.575 | 12.222 | 1.515 | 2.617 | **0.015** | 0.275 |
| **Left Precentral Gyrus** | 13.296 | 1.589 | 12.590 | 1.454 | 3.238 | **0.003** | 0.190 |
| **Right Planum Temporale** | 1.931 | 0.268 | 1.773 | 0.255 | 1.892 | 0.068 | 0.315 |
| **Left Planum Temporale** | 2.103 | 0.266 | 1.966 | 0.318 | 1.876 | 0.070 | 0.315 |
| **Right Subcallosal Area** | 1.180 | 0.115 | 1.134 | 0.140 | 1.306 | 0.169 | 0.397 |
| **Left Subcallosal Area** | 1.225 | 0.116 | 1.193 | 0.134 | 1.226 | 0.187 | 0.397 |
| **Right Superior Frontal Gyrus** | 14.897 | 1.879 | 14.673 | 1.942 | 1.876 | 0.070 | 0.315 |
| **Left Superior Frontal Gyrus** | 15.093 | 1.900 | 14.614 | 1.939 | 1.867 | 0.071 | 0.315 |
| **Right Supplementary Motor Cortex** | 5.461 | 0.713 | 5.243 | 0.712 | 2.698 | **0.012** | 0.275 |
| **Left Supplementary Motor Cortex** | 5.569 | 0.733 | 5.243 | 0.806 | 2.914 | **0.007** | 0.228 |
| **Right Supramarginal Gyrus** | 8.280 | 1.163 | 8.021 | 1.034 | 0.073 | 0.396 | 0.397 |
| **Left Supramarginal Gyrus** | 9.156 | 0.959 | 8.824 | 1.058 | 1.099 | 0.216 | 0.397 |
| **Right Superior Occipital Gyrus** | 4.173 | 0.505 | 4.072 | 0.661 | -0.779 | 0.292 | 0.397 |
| **Left Superior Occipital Gyrus** | 3.622 | 0.463 | 3.457 | 0.540 | -0.326 | 0.377 | 0.397 |
| **Right Superior Parietal Lobule** | 10.662 | 1.085 | 10.436 | 1.401 | 0.103 | 0.395 | 0.397 |
| **Left Superior Parietal Lobule** | 10.929 | 1.255 | 10.504 | 1.435 | 0.302 | 0.379 | 0.397 |
| **Right Superior Temporal Gyrus** | 7.505 | 1.002 | 7.195 | 0.893 | 0.734 | 0.303 | 0.397 |
| **Left Superior Temporal Gyrus** | 7.310 | 0.917 | 7.062 | 0.814 | 1.588 | 0.113 | 0.340 |
| **Right Temporal Pole** | 8.071 | 0.974 | 7.801 | 0.961 | 0.824 | 0.282 | 0.397 |
| **Left Temporal Pole** | 7.815 | 0.871 | 7.643 | 1.027 | 0.682 | 0.314 | 0.397 |
| **Right Triangular Part Of The Inferior Frontal Gyrus** | 3.725 | 0.488 | 3.491 | 0.524 | 2.020 | 0.053 | 0.315 |
| **Left Triangular Part Of The Inferior Frontal Gyrus** | 4.017 | 0.511 | 3.909 | 0.622 | 0.686 | 0.313 | 0.397 |
| **Right Transverse Temporal Gyrus** | 1.522 | 0.188 | 1.407 | 0.203 | 1.599 | 0.111 | 0.340 |
| **Left Transverse Temporal Gyrus** | 1.650 | 0.212 | 1.567 | 0.271 | 0.915 | 0.260 | 0.397 |
