## Supplementary figures and images for "Severe Neuro-COVID is associated with peripheral immune signatures, autoimmunity and signs of neurodegeneration: a prospective cross-sectional study"

### Figure S1

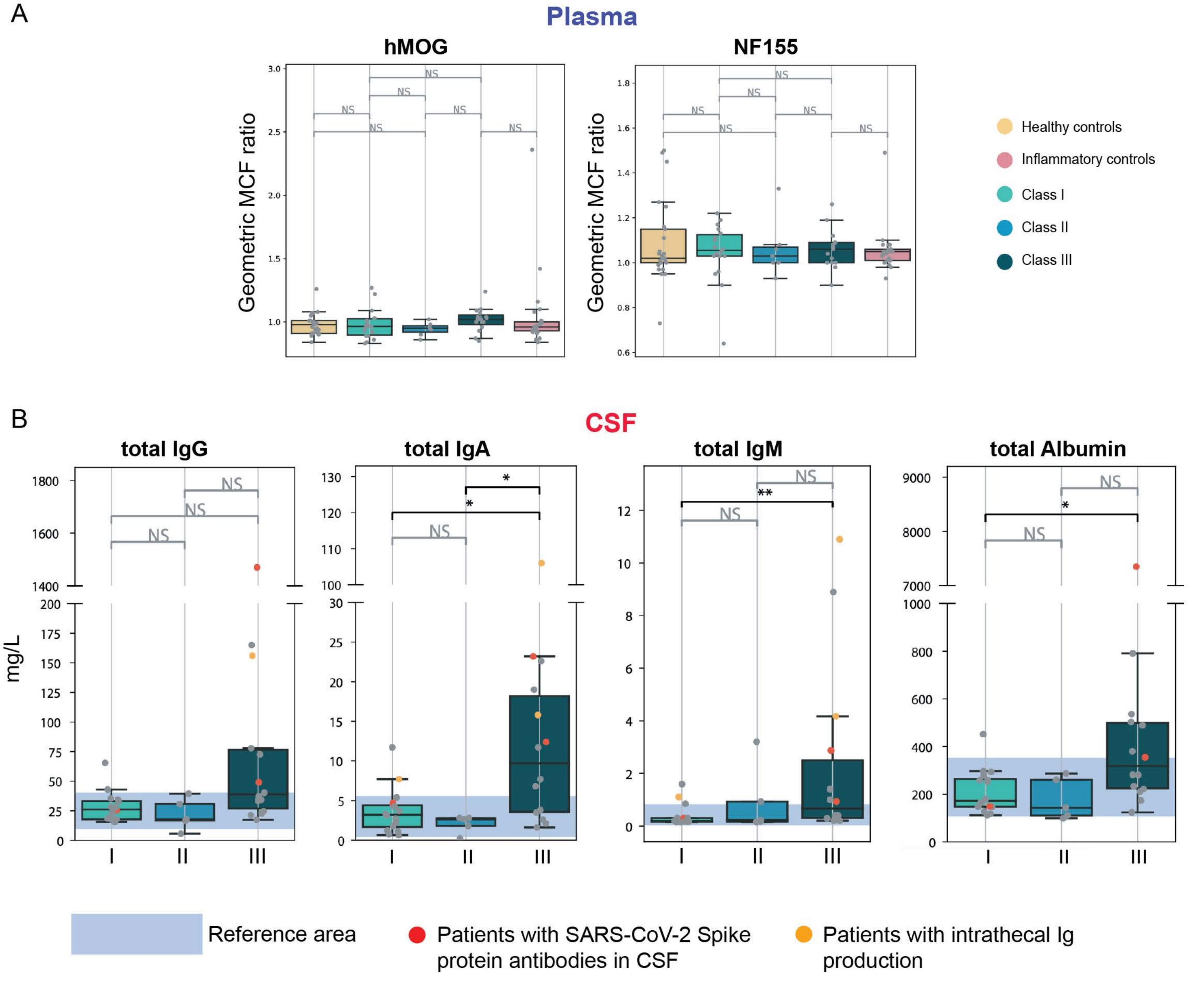

### Figure S2

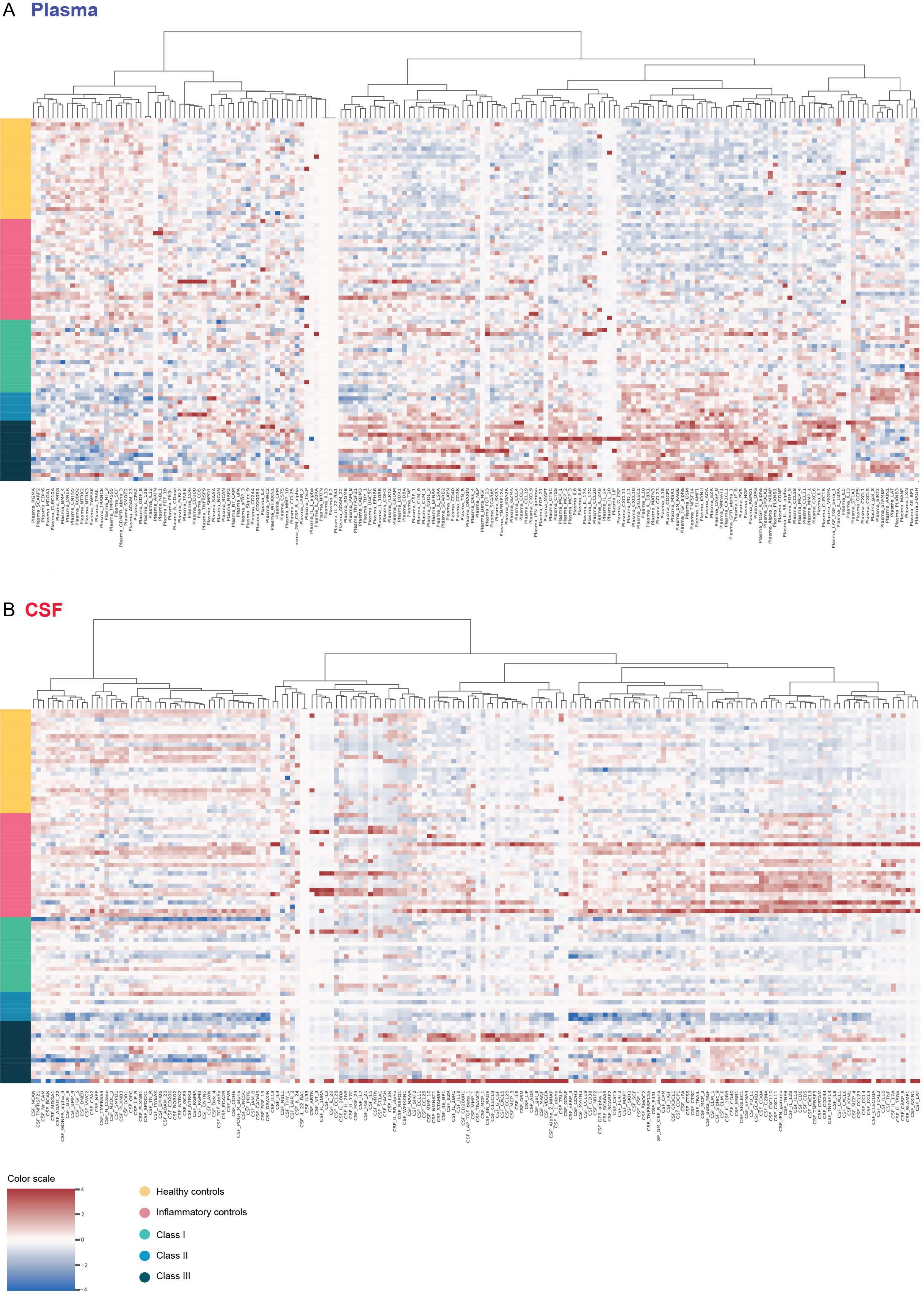

### Figure S3

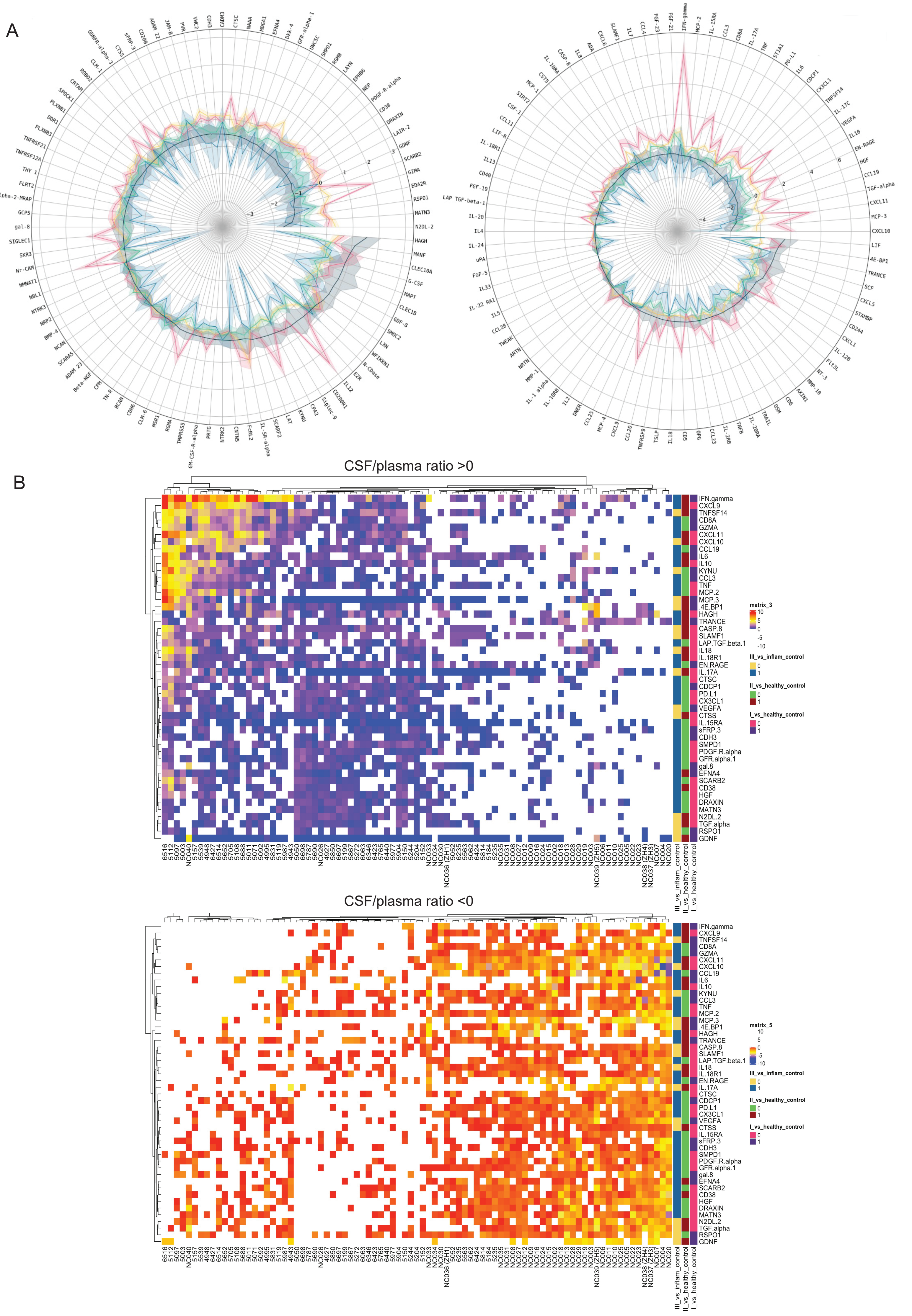

### Figure S4

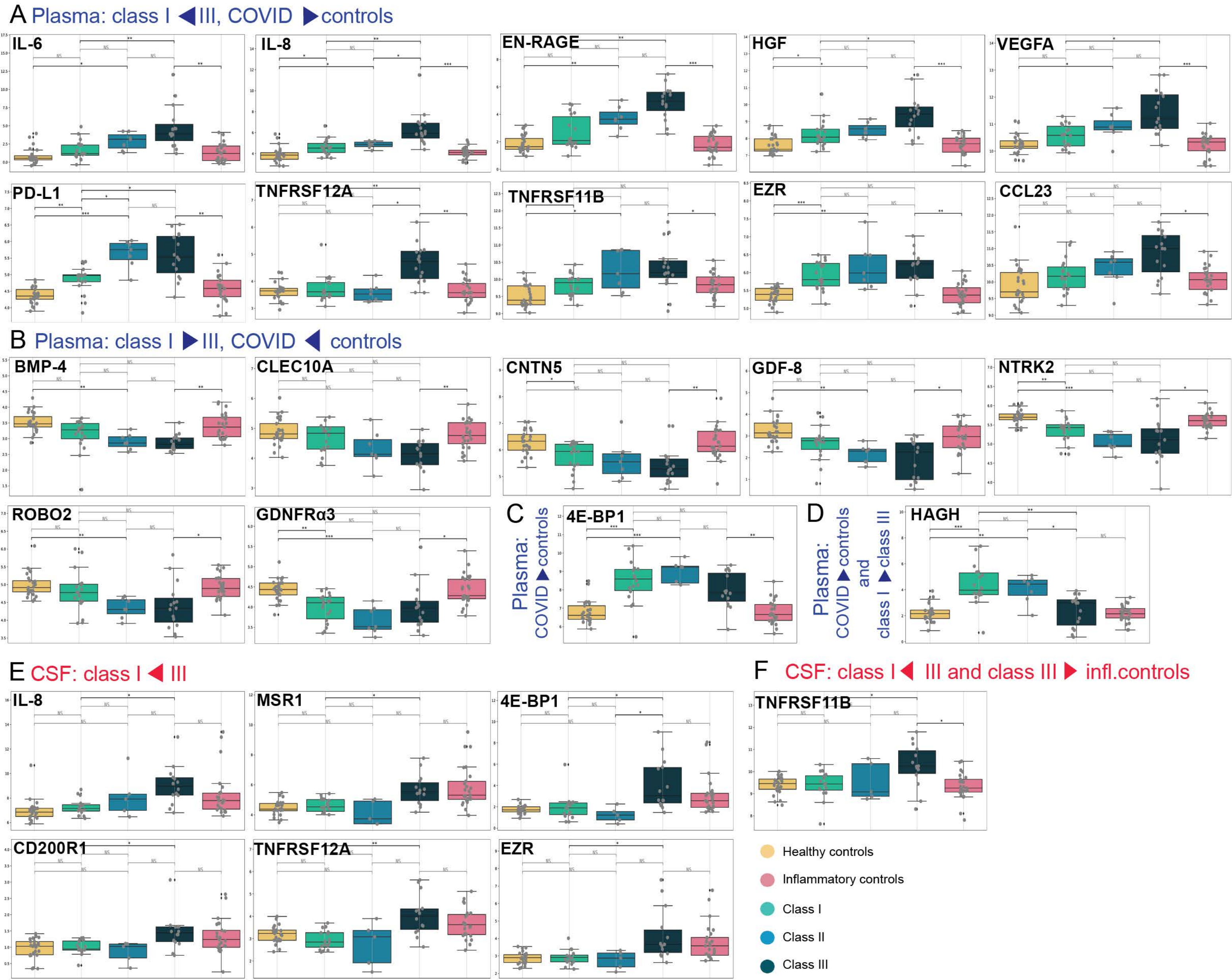

### Figure S5

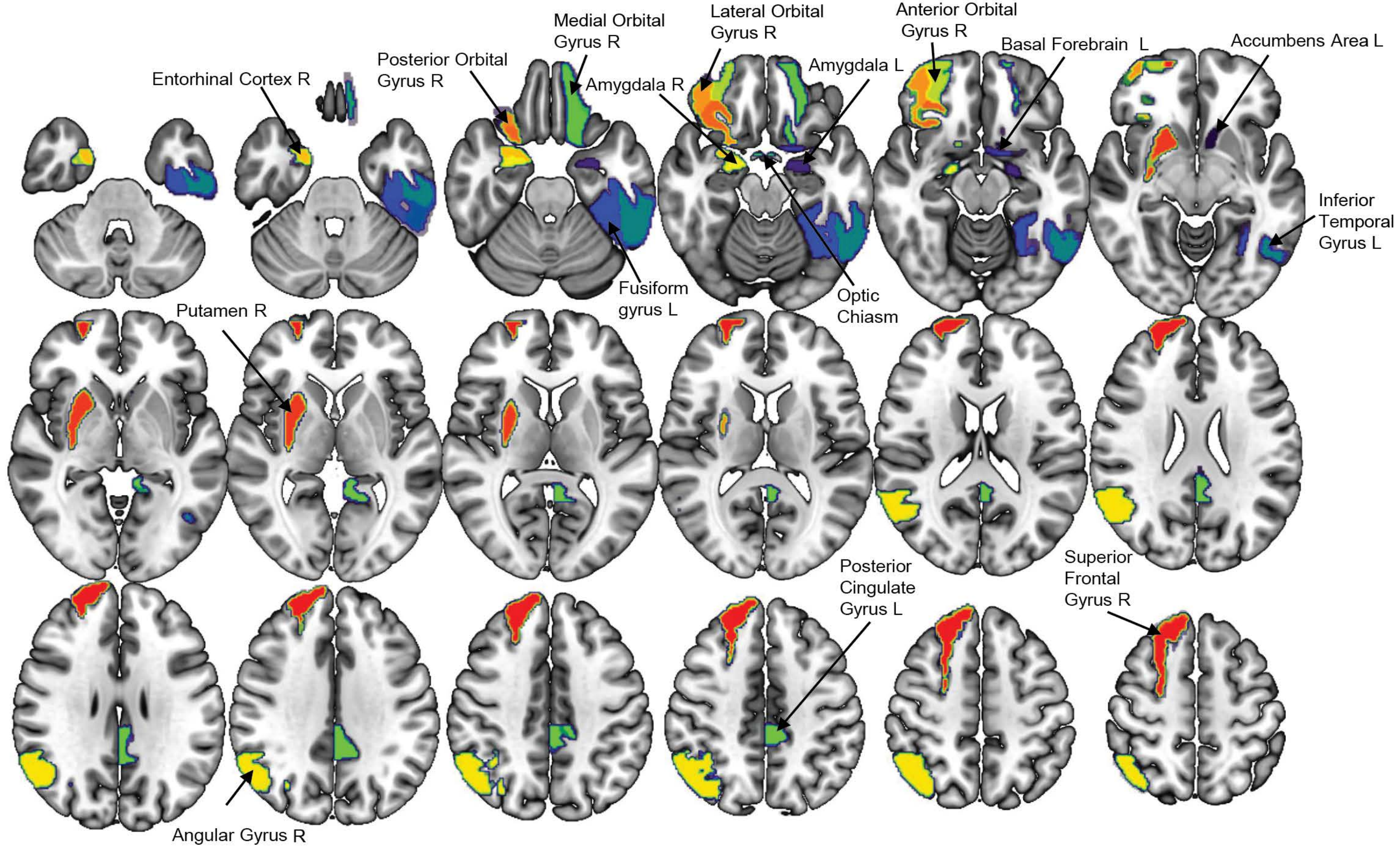

### Figure S6

A Regional brain volumes - plasma protein values

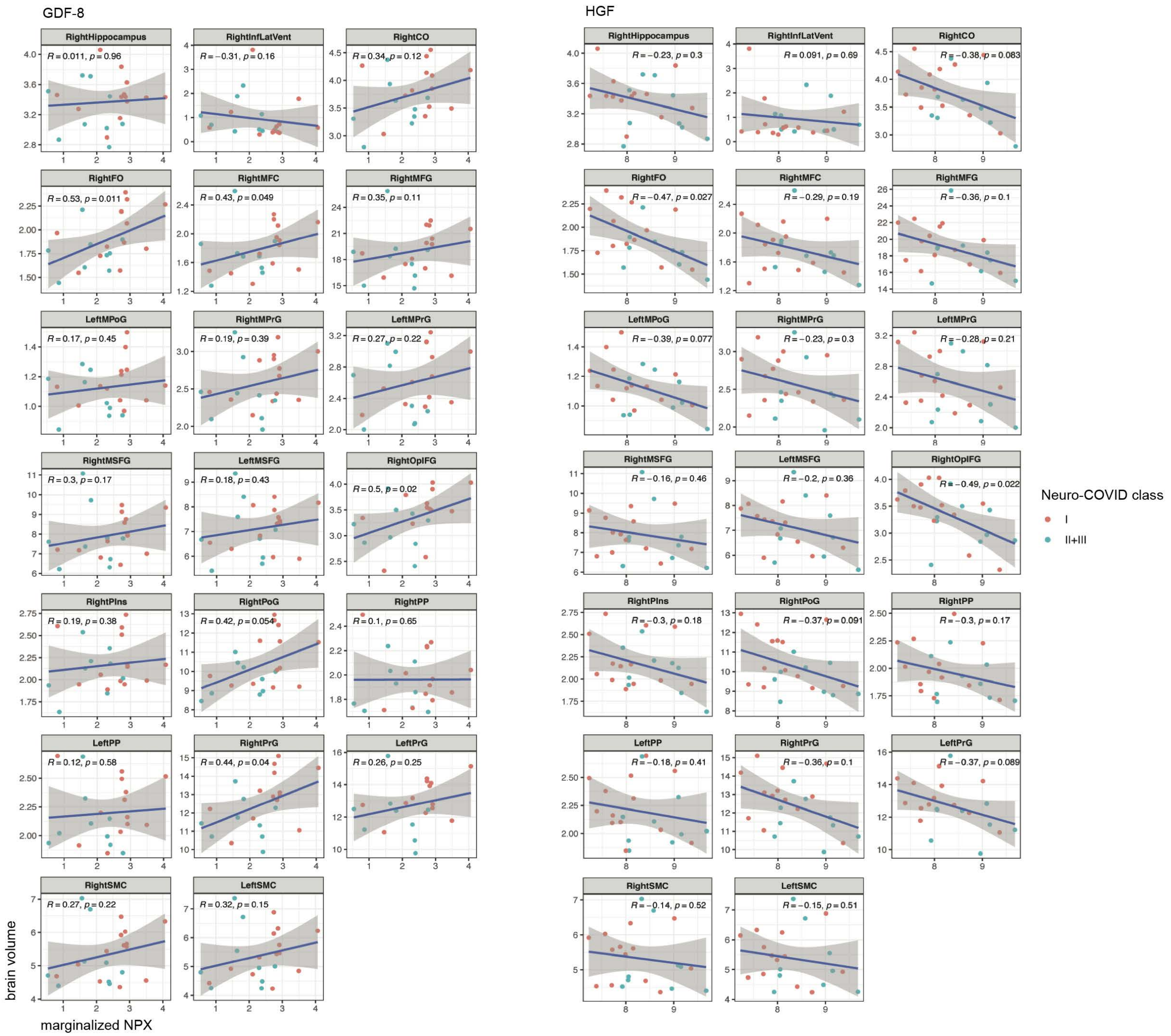


B Plasma protein values

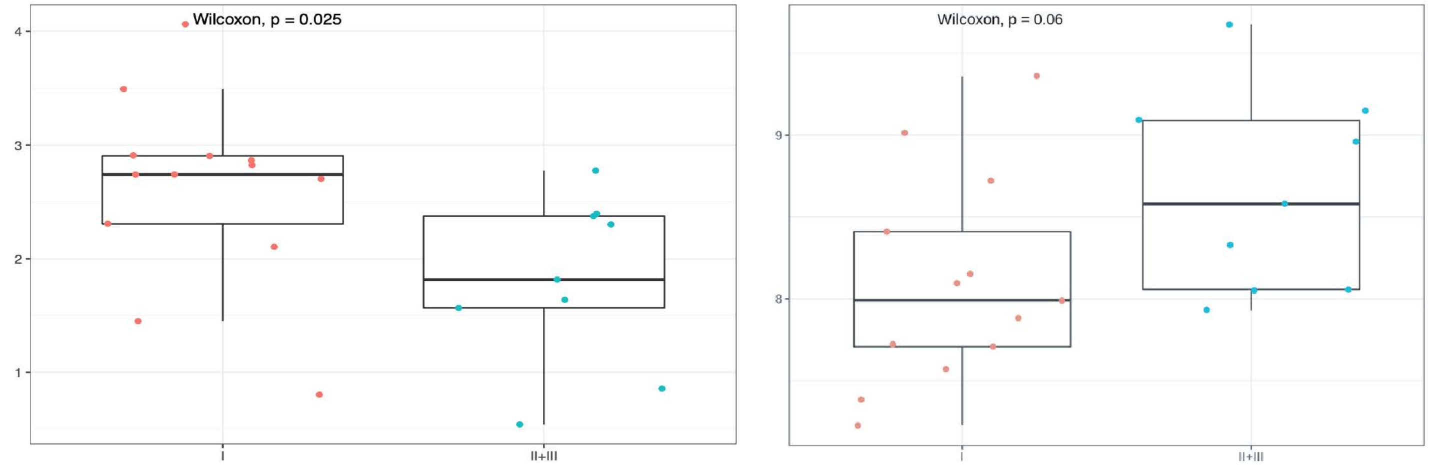

### Figure S7

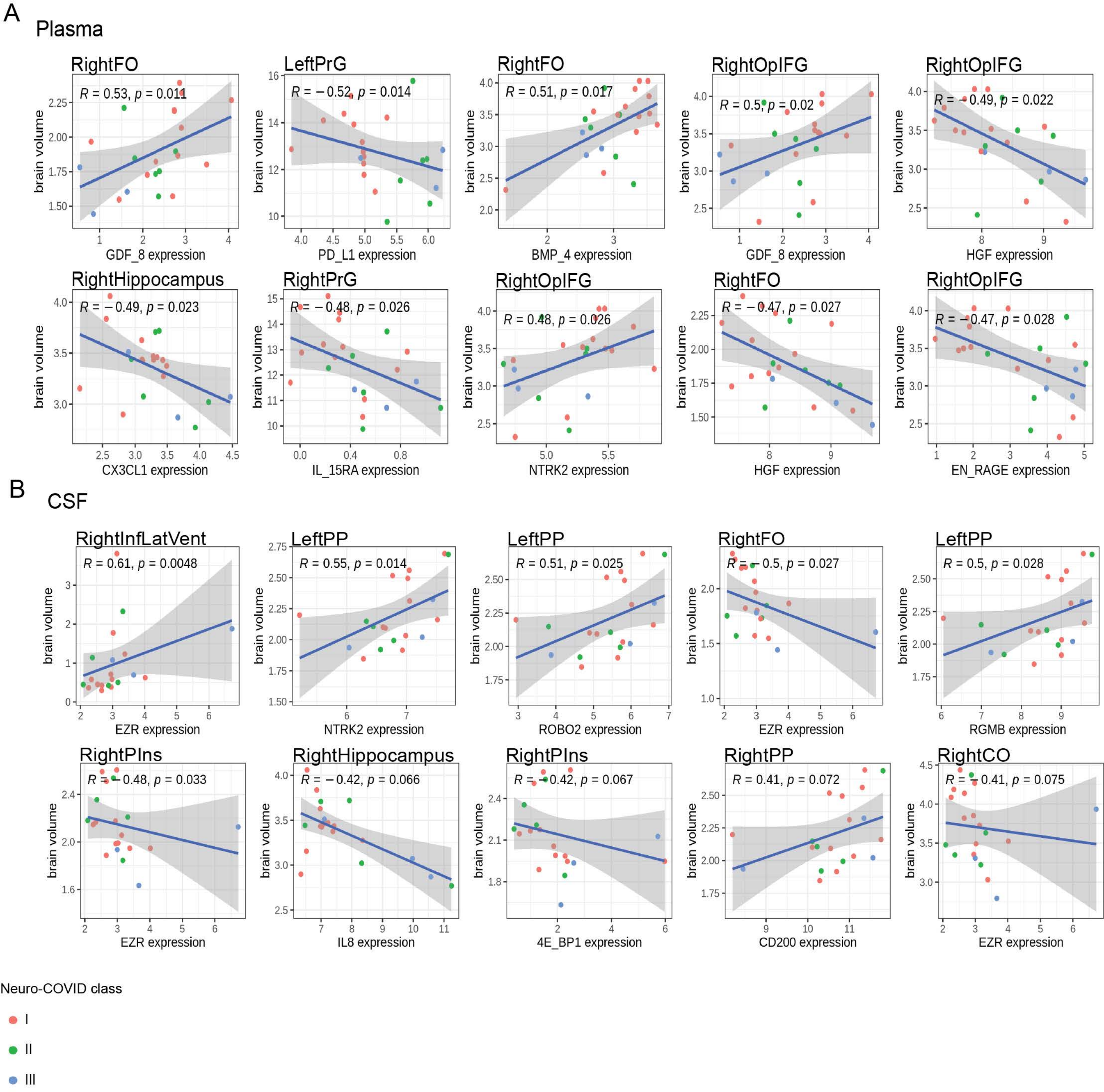

### Figure S8

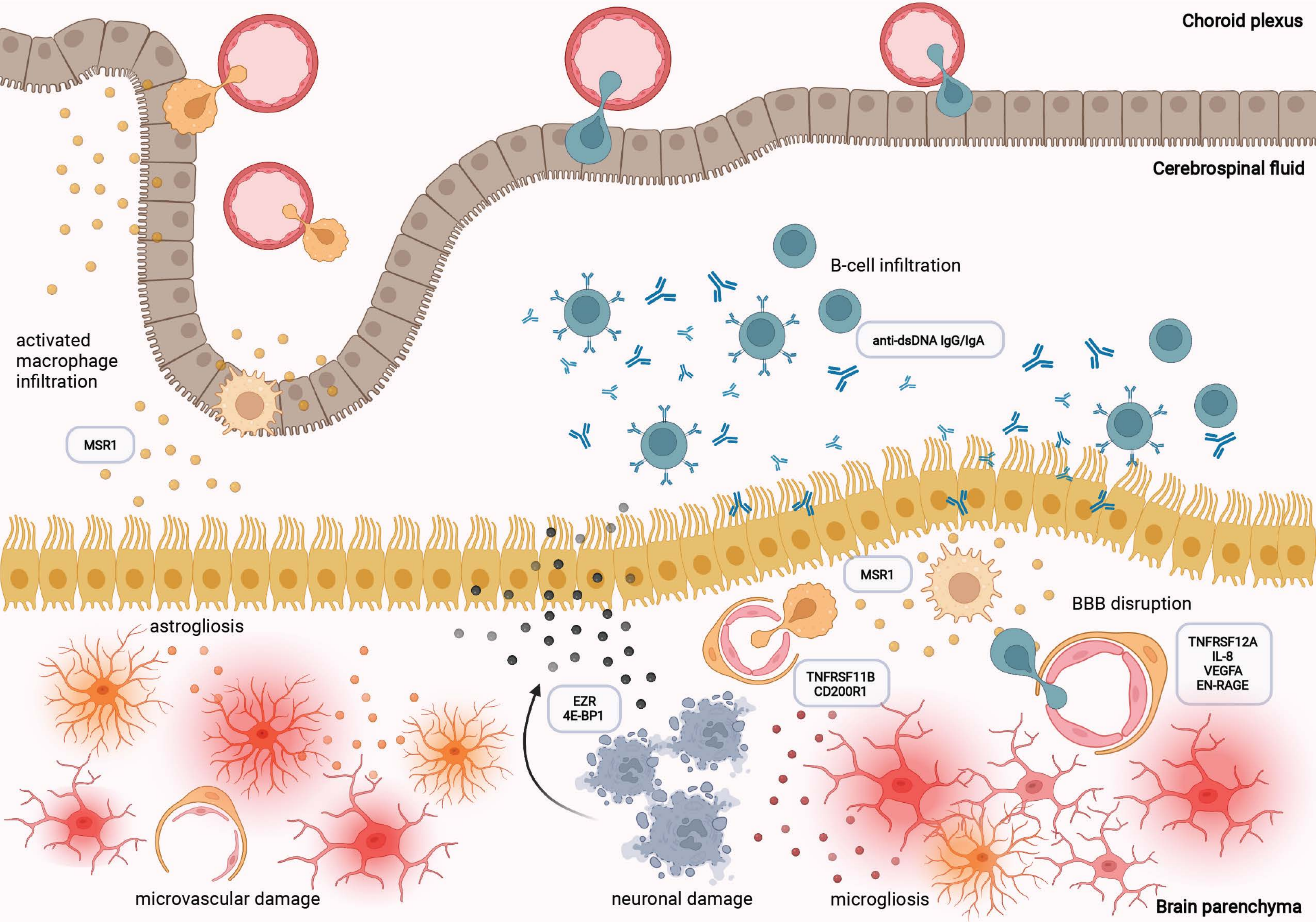
